# Secondary brain injury after parenchymal cerebral hemorrhage in humans: the role of oxidative stress and Endothelin-1

**DOI:** 10.1101/2024.09.19.24313781

**Authors:** M. De Michele, P. Amisano, O.G. Schiavo, R. Carnevale, V. Cammisotto, A. Ciacciarelli, I. Berto, U. Angeloni, S. Pugliese, D. Toni, S. Lorenzano

## Abstract

**Background and aims:** Perihematomal hypoperfusion may lead to ischemic damage during intraparenchymal cerebral hemorrhage (ICH), resulting in worse prognosis. We aimed to (1) investigate the relationship between serum biomarkers related to oxidative stress and vasoactive substances and the occurrence of hypoperfusion and ischemic perihematomal lesions in ICH, (2) to evaluate their correlation with the volumetric evolution of the hematoma and perihematomal edema.

**Methods:** We enrolled 28 patients affected by ICH. Blood samples were collected at timepoint T0 (admission to Emergency-Room), T1 (12-24hs from symptoms onset), T2 (48- 72hs from onset), to measure Endothelin-1 (ET-1), nitrites/nitrates (NO), NADPH oxidase-2 (NOX-2), metalloproteinase-12 (MMP12) and asymmetric dimethylarginine (ADMA). Patients underwent brain MRI with perfusion study at T1 and MRI without perfusion at T2.

**Results:** 12 patients had ischemic perihematomal lesions at T1. A higher NOX-2 concentration at T0 was observed in patients with ischemic perihematomal lesions compared to those without, (34.9 pg/ml vs 22.4 pg/ml, p=0.051) and with a more severe perihematomal edema at T2 (37.99 pg/ml vs. 19.17 pg/ml, p=0.011). The ischemic perihematomal lesions development was also associated with an increased hematoma volume (p<0.005), perilesional edema (p=0.046), greater midline shift (p=0.036). ET-1 values at T1 inversely correlated with hemorrhage volume at T2 (ρ=-0.717, p=0.030).

**Conclusions:** NOX-2 seems to have a role in the development of ischemic perihematomal lesions. The association between higher ET-1 values and a lower hemorrhage volume could be related to the ET-1 vasoconstriction action on the ruptured vessel wall.

## Introduction

Spontaneous intraparenchymal cerebral hemorrhage (ICH) accounts for about 20% of all strokes and is associated with high rates of mortality and long-term disability^1–2–3–4^. In recent years, besides the role of primary damage due to a direct mechanical compression by the hematoma on the surrounding tissue, many studies have highlighted the importance of the secondary damage, mostly due to the perihematomal edema and hypoperfusion, and related to oxidative stress, neuroinflammation, alteration of the blood-brain barrier (BBB), and vasoactive metabolites^5–6–7^. Hypoperfusion, together with additional factors, i.e., cerebral microangiopathy and atherosclerosis, hyperglycemia, neuroinflammation, and oxidative stress, may also be the trigger for the development of silent ischemic lesions, localized both near and in remote regions from the hematoma^8–9^. Although pathophysiology of perihematomal ischemic lesions development is not completely ascertained, the appearance of these lesions is associated with a worse prognosis and with the recurrence of cerebrovascular events (both ischemic and hemorrhagic), cognitive decline, and death^9–10–11^.

Endothelin-1 (ET-1) and Nitric oxide (NO) are two vasoactive substances with opposite action involved in autoregulation of cerebral blood flow (CBF) and neurovascular coupling^12^. ET-1 is a strong vasoconstrictor that exerts its action mainly through the ET_A_ receptors widely represented on the smooth muscle cells of brain arterioles^13^. Conversely, NO, produced by the endothelial NO synthase (NOS) isoform, acts as a powerful vasodilator^14^. There are two other NOS isoforms, one produced by neurons (nNOS) and another one produced by macrophages as response to inflammation (inducible, iNOS). nNOS-derived NO plays a critical neuroprotective role in mediating synaptic plasticity and neuronal signaling, but it becomes a neurotoxic factor when excessive amount of NO is produced. Therefore, in pathological conditions, this molecule acts as a double-edged sword^15^.

In condition of brain hypoperfusion, the pro-oxidant activity exceeds the endogenous antioxidant defense leading to oxidative stress. Similarly to oxidative stress, nitrosative stress can occur due to the excessive production of nNOS derived reactive oxygen species (ROS) and of iNOS-derived NO, which are responsible of protein, lipid and DNA damage leading to apoptosis and necrosis^16^.

Asymmetric dimethyl-arginine (ADMA) is an endogenous inhibitor of NOS and is known to be a marker of endothelial disfunction and atherosclerosis. It seems to be involved in CBF reduction and in promoting oxidative stress and inflammatory response to ischemia^17^.

Nicotinamide adenine dinucleotide phosphate (NADPH) oxidase (NOX) is a complex multimeric enzyme that participates in the generation of superoxide or hydrogen peroxide^18–19^. NOX2 is the most represented isoform in cerebral arteries and is involved in the oxidative stress-related damage^20^.

Matrix metalloproteinases (MMP) are a superfamily of calcium-dependent zinc-containing endopeptidases with a wide range of biological functions^21^. MMP12 is a metalloelastase produced by macrophages which exerts a role in post-ischemic brain injury contributing to blood- brain barrier (BBB) break-down, inflammation, apoptosis and demyelination^22,23^.

Main aim of this study was to assess the role of vasoactive substances (ET1 and NO), oxidative stress markers (ADMA and NOX2) and neuroinflammation mediators (MMP12) in the development of perihematomal hypoperfusion and ischemic lesions. Secondly, we aimed to study the correlation between these molecular biomarkers and the evolution of hematoma and perilesional edema volume. Finally, we evaluated the impact of these variables on clinical outcome.

## Materials and methods

### Patients

In this study, we included patients with spontaneous ICH, admitted to the Emergency Department of our hospital, within 6 hours of symptom onset, between January 2019 and July 2021. For each patient we collected demographic data, pre-stroke modified Rankin Scale (mRS), vascular risk factors, previous medical therapy, presence of infections in the previous two weeks, stressful factors at the time of the event. We defined three time points from symptom onset for the collection of clinical and diagnostic data: time 0, at admission (T0); time 1 (T1) at 12-24h and time 2 (T2) at 48-72h. At T0, we performed general and neurological examination, measurement of vital parameters (blood pressure, heart rate and body temperature) and of serum glucose, assessment of stroke severity by using the National Institutes of Health Stroke Scale (NIHSS) score and plain brain Computed Tomography (CT) scan. We took blood samples at the three prespecified timepoints for the routine blood tests and for dosing the following molecular biomarkers: ET-1, NO, NOX2, ADMA, and MMP12. The T0 sampling was collected prior to the administration of any drug. At T1, we performed a brain Magnetic Resonance Imaging (MRI) including diffusion (DW) and perfusion (PW) weighted images, clinical evaluation with NIHSS and blood pressure measurement. At T2, we undertook a follow-up MRI (DWI, ADC, Fluid attenuated inversion recovery (FLAIR) and Gradient Echo (GE).

Finally, we evaluated the level of disability at 3 months after the acute event using the modified Rankin scale (mRS) by telephone interview (T3).

A schematic representation of the study experimental design is depicted in Figure 1.

**Figure 1.**
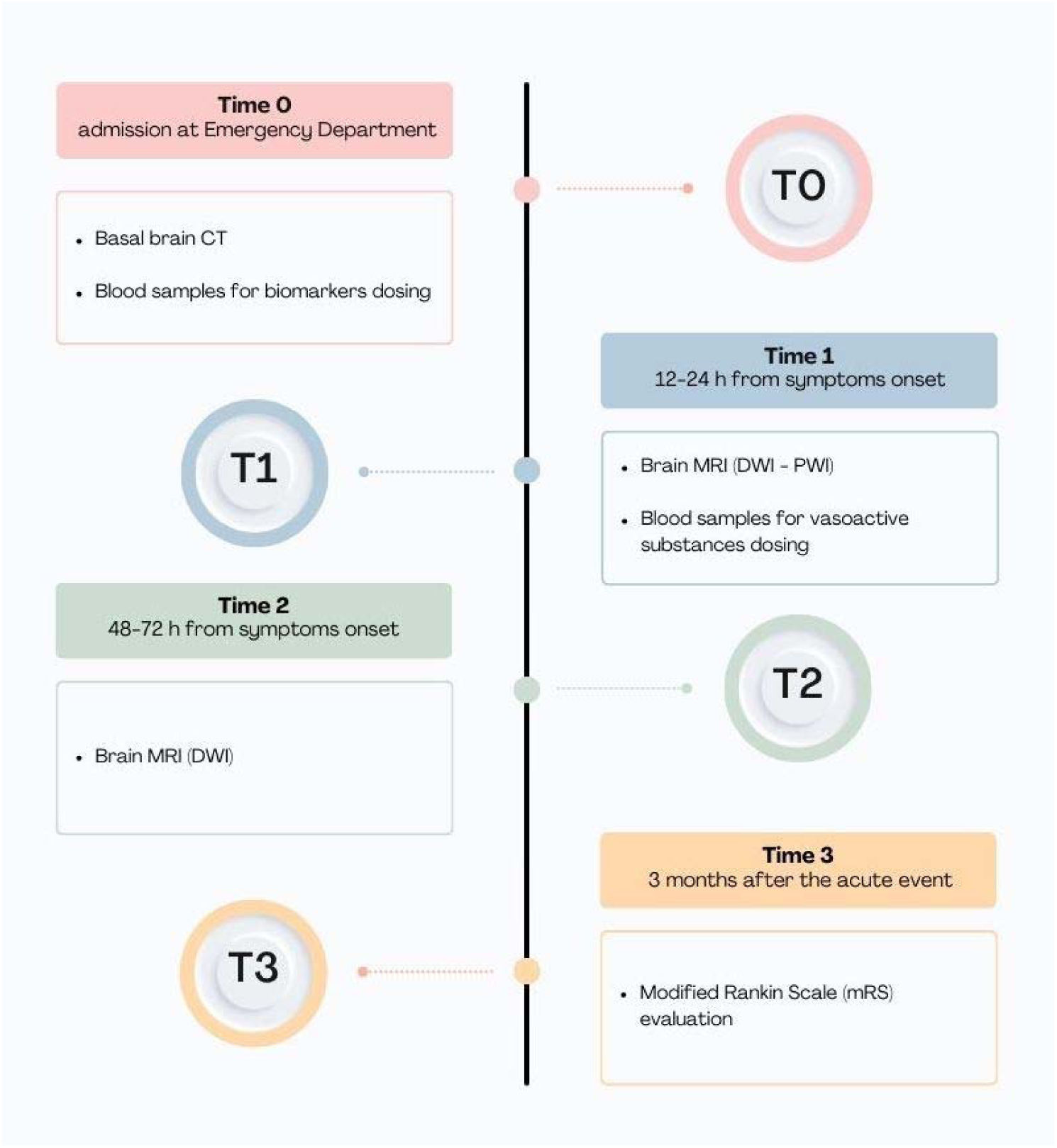
Study timeline.

All study participants gave a written informed consent. The study was approved by the Ethical Committee of the Policlinico Umberto I University Hospital (number 5875) and was conducted according to the Declaration of Helsinki.

### Laboratory data

Blood samples were centrifuged; serum was aliquoted and stored at – 80°C until the time of analysis. The dosages of ET-1, Nitrite/nitrates (metabolism end products of NO and then NO surrogate markers), NOX2, ADMA and MMP12 were carried out with enzyme-linked immunoassay (ELISA) method.

### Radiological data

Two independent investigators used the ABC/2 formula to calculate the hematoma volume. At CT images, we applied the ABC/2 formula to the hemorrhagic lesion visible as hyperdensity (+ 30-45 Hounsfield unit, HU). At MRI images, we applied the formula to the visible hypointensity in the GRE sequence, while for the calculation of the edema volume the score was applied to the visible hyperintensity in the FLAIR sequence. Perihematomal edema volume was measured by subtracting the hematoma volume from the combined hematoma and perihematomal hyperintensity area volumes. Perihematomal edema was considered mild if only sulci effacement was present, moderate in the presence of ventricle asymmetry, and severe if there was midline shift. The midline shift (mm) was also measured. We also took into consideration the hematoma growth defined as any volume change from T1 to T2. The “island sign”, defined as ≥3 small discrete hematomas that are not connected to the primary hematoma or ≥4 small hematomas that may branch off from the primary hematoma and resemble “bubbles” or “sprouts” rather than a lobulated appearance, was evaluated at the basal brain CT. Finally, we considered Mean Transit Time (MTT), Cerebral Blood Flow (CBF), and Cerebral Blood Volume (CBV), for the evaluation of hypoperfusion related to the hemorrhagic lesion. We measured CBF, CBV, and MTT values in 5 round regions of interest of about 0.5 cm^2^ within 1 cm from the hematoma periphery and drawn freehand on the slice in which the hemorrhage was more visible. Values were averaged and expressed as continuous and categorical variables. According to previously published paper^6^, CBF was categorized into normal (40–55 mL/100 g/min) and low (<40 mL/100 g/min); CBV was dichotomized into normal (>2.5 mL/100 g) and low (≤2.5 mL/100 g) and MTT was dichotomized into normal (≤5 s) and high (>5 s).

According to the results from MRI images, we divided the patients in two groups: patients with the presence of one or more ischemic lesions in the diffusion sequences at T1 and patients without ischemic lesions at T1.

### Statistical analysis

The descriptive statistical analysis of the total population and of the two groups of interest was carried out by calculating means or medians for continuous variables based on their normal or non-normal distribution as well as by calculating frequencies, proportions (or percentages) for dichotomic/categorical variables. For comparison between the two groups of patients with ischemic lesions and patients without ischemic lesions: continuous variables with normal and non-normal distributions were compared using the t-test for paired samples or the Mann-Whitney U-test, respectively. We compared categorical variables using the Fisher test or chi-square test, when appropriate. Correlations between variables were assessed by calculating the Pearson or Spearman coefficient. We presented the course of the molecular biomarkers across the different time points, as well as the median changes over time by presence/absence of ischemic lesions at T1. Given the small numbers of patients included in the study, we performed an exploratory multivariate logistic regression to detect potential predictors of the development of perihematomal ischemic lesions after adjustment for age, sex and variables with a univariate p value <0.05. For all tests that were performed, a value of p<0.05 was considered statistically significant. Statistical analysis was performed using SPSS statistical software (IBM Corp, SPSS Statistics for Windows, Version 25, Armonk, NY).

## Results

We enrolled 28 patients with ICH, 15 males (55.2%) and 13 females (44.8%), mean (± SD) age 70.2 years (± 13.8). Pre-stroke mRS was between 0 and 1 in 24 (85.2%) patients. Overall, 12 patients (42.8%) had one or more ischemic lesions in DWMRI at T1, while there were 16 patients (57.1%) without ischemic lesions.

### Patients’ characteristics

We did not find any statistically significant differences between the two groups as regards demographic characteristics and NIHSS score, blood pressure, heart rate and serum glucose values at the three assessment timepoints (Table 1 and Table 2). However, we found a higher prevalence of past medical history of hypertension (12 [100%] vs 9 [56.3%] patients; p= 0.010) and of dyslipidemia (9 [75%] patients vs 1 [6.3%] patient, p<0.001) and a higher prevalence of neutrophil leukocyte counts at T0 (7.44 ×10^3^/µL vs 4.99 ×10^3^/µL; p <0.037) in the group of patients with ischemic lesions.

**Table 1.** Clinical and demographic characteristics of patients according to presence/absence of ischemic lesions at T1.

|  | **All patients**<br>**N=28** | **Ischemic lesions**<br>**n=12** | **No ischemic lesions**<br>**n=16** | **p** |
| --- | --- | --- | --- | --- |
| ***Demographic characteristics and clinical history*** |  |  |  |  |
| Age, mean (SD) | 70.2 (13.8) | 68.4 (15.5) | 70.8 (13.0) | 0.661 |
| Sex (female) (%) | 13 (44.8) | 6 (50) | 6 (37.5) | 0.508 |
| Pre-stroke mRS (%) |  |  |  | 0.620 |
| - 0 | 19/28 (67.9) | 7/11 (63.6) | 12 (75.0) |  |
| - 1 | 5/28 (17.9) | 2/11 (18.2) | 3 (18.8) |  |
| - 2 | 1/28 (3.6) | 0 | 0 |  |
| - 3 | 3/28 (10.7) | 2/11 (18.2) | 1 (6.3) |  |
| - 4 | 0 | 0 | 0 |  |
| - 5 | 0 | 0 | 0 |  |
| Pre-stroke mRS 0-1 (%) | 23/27 (85.2) | 9/11 (81.8) | 14/15 (93.3) | 0.556 |
| Smoking (%) |  |  |  | 0.174 |
| - no | 7 (24.1) | 3 (25.0) | 4 (25.0) |  |
| - current | 15 (51.7) | 8 (66.7) | 6 (37.5) |  |
| - previous | 7 (24.1) | 1 (8.3) | 6 (37.5) |  |
| Obesity (%) | 7 (24.1) | 3 (25.0) | 4 (25.0) | 1.0 |
| Alcohol consumption (%) | 2 (6.9) | 0 | 2 (12.5) | 0.492 |
| Abuse drug consumption (%) | 1 (3.4) | 0 | 1 (6.3) | 1.0 |
| Hypertension (%) | 22 (75.9) | 12 (100.0) | 9 (56.3) | **0.010** |
| Dyslipidemia (%) | 10 (34.5) | 9 (75.0) | 1 (6.3) | **<0.001** |
| Atrial fibrillation (%) | 6 (20.7) | 2 (16.7) | 3 (18.8) | 1.0 |
| Ischemic cardiopathy (%) | 4 (13.8) | 1 (8.3) | 3 (18.8) | 0.613 |
| Diabetes Mellitus (%) | 7 (24.1) | 4 (33.3) | 3 (18.8) | 0.418 |
| Previous stroke (%) | 2 (6.9) | 0 | 2 (12.5) | 0.492 |
| Previous TIA (%) | 0 | 0 | 0 | - |
| Coagulopathy (%) | 1 (3.4) | 0 | 1 (6.3) | 1.0 |
| Valvulopathy (%) | 2 (6.9) | 0 | 1 (6.3) | 1.0 |
| Peripheral vasculopathy (%) | 1 (3.4) | 0 | 1 (6.3) | 1.0 |
| Migraine (%) | 0 | 0 | 0 | - |
| Cancer (%) | 2 (6.9) | 1 (8.3) | 1 (6.3) | 1.0 |
| Stressful events (%) | 3 (10.3) | 1 (8.3) | 2 (12.5) | 1.0 |
| ***Therapy before hemorrhagic stroke*** |  |  |  |  |
| Anticoagulants (%) | 7 (24.1) | 2 (16.7) | 4 (25.0) | 0.673 |
| Antiplatelets (%) | 8 (28.6) | 4/11 (36.4) | 4 (25.0) | 0.675 |
| Lipid lowering medications (%) | 2 (6.9) | 2 (16.7) | 0 | 0.175 |
| ACE-I/ARB (%) | 12 (42.9) | 7/11 (63.6) | 4 (25.0) | 0.061 |
| Diuretics (%) | 5 (17.2) | 2 (16.7) | 2 (12.5) | 1.0 |
| Beta-blockers (%) | 5 (17.2) | 3 (25.0) | 2 (12.5) | 0.624 |
| Alpha-lytics (%) | 4 (13.8) | 3 (25.0) | 1 (6.3) | 0.285 |
| Calcium-antagonists (%) | 0 | 0 | 0 | - |
| PPI (%) | 3 (10.3) | 1 (8.3) | 2 (12.5) | 1.0 |
| Blood glucose lowering medications (%) | 3 (10.3) | 2 (16.7) | 1 (6.3) | 0.560 |
| Insulin (%) | 0 | 0 | 0 | - |
| Nitrates (%) | 0 | 0 | 0 | - |
| ***Clinical characteristics of hemorrhagic stroke*** |  |  |  |  |
| Stroke on awakening (%) | 11/29 (37.9) | 7 (58.4) | 4 (25.0) | 0.121 |
| NIHSS, median (IQR) |  |  |  |  |
| - Admission (T0) | 11 (7.5-18) | 14 (6.75-17.50) | 9 (7.25-18.25) | 0.642 |
| - 12-24 h (T1) | 9 (6.5-13.5) | 13 (6-20) | 9 (7.75-10.50) | 0.667 |
| - 48-72 h (T2) | 10 (7.5-14.5) | 14 (6-20) | 8.50 (7.75-10.50) | 0.197 |
| - Discharge | 7 (5-11.5) | 8 (4-12.50) | 6 (5-11) | 0.720 |
| SBP, mean (SD) |  |  |  |  |
| - T0 | 168.4 (31.50) | 168.5 (30.3) | 168.9 (34.4) | 0.977 |
| - T1 | 145.8 (25.9) | 150.1 (24.3) | 140.7 (29.0) | 0.534 |
| - T2 | 147.2 (22.4) | 149.9 (13.9) | 143.4 (32.7) | 0.646 |
| DBP, mean (SD) |  |  |  |  |
| - T0 | 92.57 (19.21) | 91.4 (20.1) | 94.0 (19.7) | 0.740 |
| - T1 | 85.8 (17.6) | 88.9 (16.1) | 81 (19.4) | 0.387 |
| - T2 | 84.1 (16.7) | 83.3 (10.0) | 85.2 (24.8) | 0.855 |
| HR, mean (SD) |  |  |  |  |
| - T0 | 80.6 (13.71) | 78.6 (15.5) | 81.1 (12.6) | 0.652 |
| - T1 | 81.2 (21.2) | 88 (22.6) | 73.3 (18.1) | 0.228 |
| - T2 | 79.3 (12.3) | 83.9 (19.6) | 73.0 (10.0) | 0.362 |
| Body temperature, mean (SD) |  |  |  |  |
| - T0 | 36.0 (0) | 36 (0) | 36.0 (0) | - |
| - T1 | 36.9 (0.70) | 27.2 (0.8) | 36.6 (0.3) | 0.108 |
| - T2 | 36.9 (0.6) | 37.2 (0.4) | 36.6 (0.6) | **0.043** |
| HGT, mean (SD) |  |  |  |  |
| - T0 | 128 (29.4) | 136 (32.9) | 120 (24.6) | 0.143 |
| - T1 | 128 (32.7) | 128 (30.9) | 130 (38.7) | 0.949 |
| - T2 | 119 (23.0) | 118 (25.8) | 120 (97.5-137.5) | 0.900 |
| Craniotomy (%) | 3/19 (15.8) | 2/8 (25.0) | 1/11 (9.1) | 0.546 |

SD=standard deviation; TIA=transient ischemic attack; ACE-I/ARBs= angiotensin converting enzyme inhibitors/angiotensin II receptor blockers; PPIs= Proton-Pump Inhibitors; NIHSS= National Institutes of health Stroke Scale; SBP= systolic blood pressure; DBP= diastolic blood pressure; HR= heart rate; HGT= Hemo Glucose Test

**Table 2.** Blood test results of patients according to presence/absence of ischemic lesions at T1.

|  | **All patients**<br>**N=28** | **Ischemic lesions**<br>**n=12** | **No ischemic lesions**<br>**n=16** | **p** |
| --- | --- | --- | --- | --- |
| ***Routine blood tests*** |  |  |  |  |
| Time onset-blood sampling T0, h, median (IQR) | 2.08 (1.27-2.57) | 2.33 (1.24-4.19) | 2.07 (1.14-2.41) | 0.429 |
| Time onset-T1, h, median (IQR) | 26.50 (25.30-27.30) | 26.5 (25.6-29.5) | 26.60 (25.10, 27.58) | 0.927 |
| Time onset-T2, h median (IQR) | 51.0 (48.5-53.2) | 51 (50.25-54.60) | 49.9 (48.3-52.9) | 0.361 |
| Haematocrit, T0, mean (SD) | 40.5 (5.1) | 38.7 (5.2) | 42.2 (4.7) | 0.073 |
| Platelets, T0, mean (SD) | 232.5 (74.95) | 233.3 (67.3) | 222.8 (75.2) | 0.703 |
| **Total leukocytes, mean (SD)** |  |  |  |  |
| - T0 | 8.66 (2.96) | 10.18 (3.31) | 7.42 (2.17) | **0.013** |
| - T1 | 10.51 (5.24) | 12.32 (5.90) | 7.50 (2.21) | 0.234 |
| - T2 | 11.83 (7.87) | 13.52 (9.40) | 8.46 (1.32) | 0.400 |
| **Neutrophils (absolute number), mean (SD)** |  |  |  |  |
| - T0 | 6.13 (2.91) | 7.44 (3.26) | 4.49 (2.35) | **0.037** |
| - T1 | 8.46 (5.11) | 10.2 (5.83) | 5.64 (2.06) | 0.254 |
| - T2 | 9.94 (7.36) | 11.55 (8.76) | 6.71 (1.14) | 0.387 |
| **Neutrophils (%)** |  |  |  |  |
| - T0 | 68.90 (12.94) | 71.20 (11.88) | 67.4 (14.2) | 0.456 |
| - T1 | 77.56 (8.06) | 79.62 (9.25) | 74.13 (5.26) | 0.392 |
| - T2 | 81.86 (5.26) | 83.23 (6.03) | 79.10 (1.66) | 0.295 |
| **Lymphocytes (absolute number), mean (SD)** |  |  |  |  |
| - T0 | 1.84 (1.03) | 1.94 (1.0) | 1.70 (1.08) | 0.555 |
| - T1 | 77.56 (8.06) | 1.31 (0.62) | 1.11 (0.19) | 0.612 |
| - T2 | 1.08 (0.35) | 1.09 (0.43) | 1.06 (0.16) | 0.912 |
| **Lymphocytes (%), mean (SD)** |  |  |  |  |
| - T0 | 22.40 (11.35) | 20.24 (9.71) | 23.69 (12.81) | 0.443 |
| - T1 | 13.60 (5.95) | 12.68 (7.52) | 15.13 (2.29) | 0.612 |
| - T2 | 10.41 (3.20) | 9.23 (3.36) | 12.77 (0.56) | 0.123 |
| **PCR, mean (SD)** |  |  |  |  |
| - T0 | 0.51 (0.72) | 0.79 (0.97) | 0.23 (0.24) | 0.111 |
| - T1 | 2.33 (1.68) | 3.10 (1.59) | 1.06 (0.95) | 0.094 |
| - T2 | 6.17 (6.19) | 8.70 (6.75) | 1.94 (0.07) | 0.144 |
| **INR, T0, mean (SD)** | 1.09 (0.22) | 1.04 (0.10) | 1.13 (0.28) | 0.253 |
| **PTT ratio, T0, mean (SD)** | 0.96 (0.23) | 0.94 (1.76) | 0.97 (0.27) | 0.804 |
| **Fibrinogen, T0, mean (SD)** | 374.38 (90.50) | 389.3 (84.5) | 358.81 (96.37) | 0.391 |
| **Total cholesterol, T0, mean (SD)** | 162.63 (32.02) | 161.0 (21.6) | 164.3 (43.8) | 0.898 |
| **LDL, T0, mean (SD)** | 103.25 (23.40) | 109.5 (19.7) | 97.0 (28.0) | 0.493 |

IQR= Interquartile range; PCR= Polymerase Chain Reaction; INR= International Normalized Ratio; PTT= Prothrombin Time Test; LDL= Low Density Lipoproteins

### Radiological features

Patients with ischemic lesions (Figure 2) had a median hematoma volume at T1 significantly larger than that of patients without ischemic lesions (23.05 cm3 vs 6.65 cm3, p<0.005) and a significantly higher volume of perilesional edema (15.25 vs 8.25 cm3, p 0.046) with a more severe midline shift [4.45 mm (±4.74) vs 1.31 mm (±2.62) (p=0.036)] (Supplemental Table 1). This data was also confirmed at T2 (Supplemental Table 1). The “island sign” was more frequently observed in group 1 (10 [83.3%] vs 4/15 patients [26.7%], p 0.003). When associations between prespecified perfusion parameters and the development of ischemic lesions at T1 or T2 were investigated, we did not find statistically significant differences.

**Figure 2.**
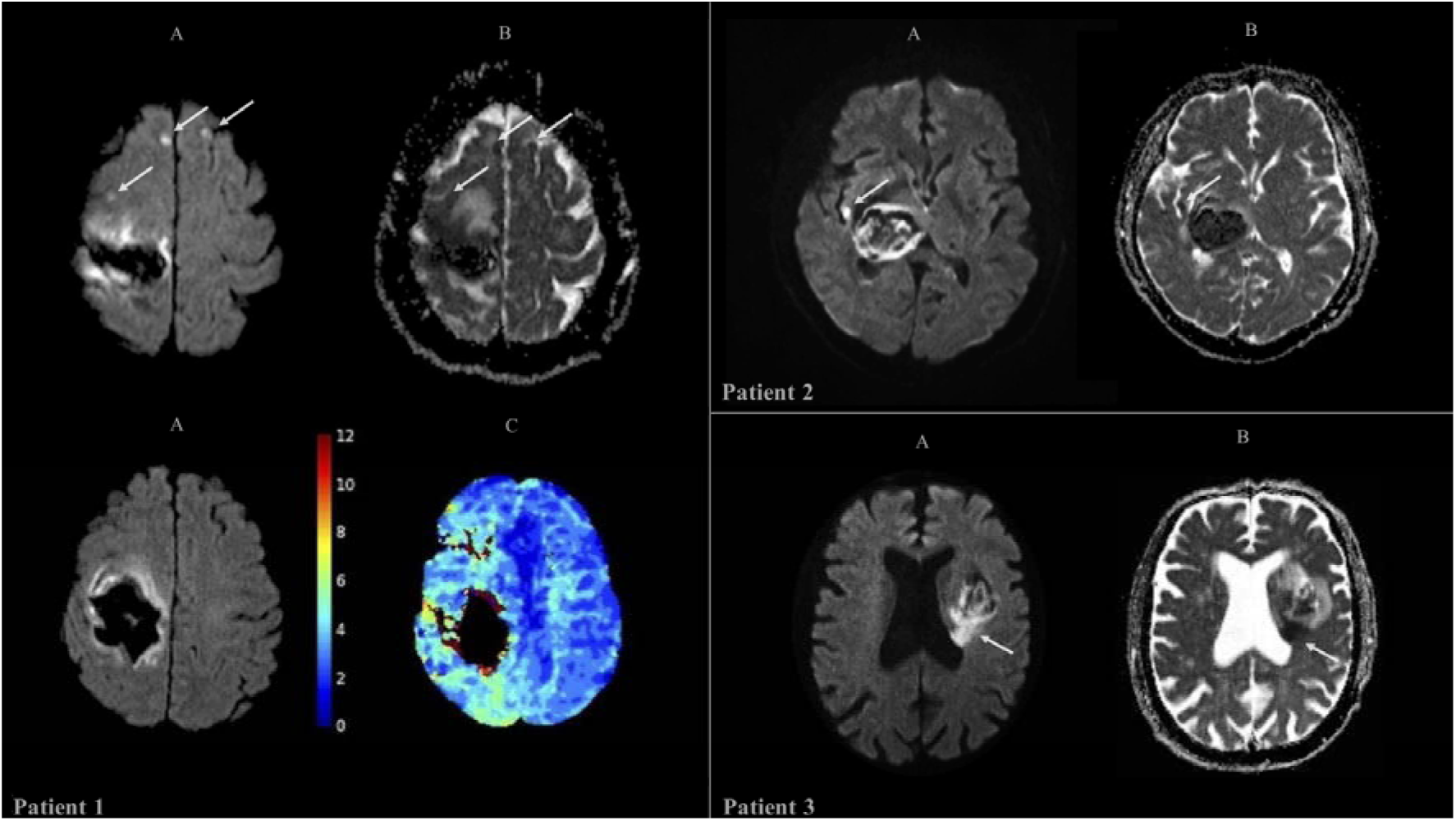
Representative neuroimaging of three patients. Patient 1. MRI imaging at T2 reveals a large intraparenchymal hematoma within the right fronto-parietal lobe. (**A**) DWI sequences identify linear regions of restricted diffusion in the anterior perilesional area and the posteromedial region, with corresponding hypointensity on ADC map (**B**), suggestive of ischemic injury. Additionally, small ischemic foci, measuring a few millimeters, are observed in the right frontal subcortical paramedian region and at the left frontal subcortical level. (**C**) Perfusion-weighted imaging (PWI) shows a significant increase of Tmax within the right parieto-occipital region and posterior temporal lobe. Patient 2. MRI imaging at T1 demonstrates a hematoma located in the right thalamo-capsular region with slight contralateral shift of the midline structures. (**A**) DWI imaging shows a perihematomal 5 mm^2^ area of diffusion restriction, with corresponding hypointensity on the ADC map (**B**), consistent with a small recent ischemic lesion. Patient 3. MRI imaging at T1 reveals a hemorrhagic focus in the left insula and basal ganglia, with slight lateral ventricle compression. (**A**) DWI imaging demonstrates a region of restricted diffusion in the posteromedial perilesional area, with corresponding hypointensity on ADC map (**B**), suggestive of ischemic injury.

### Clinical outcomes

No statistically significant between-group differences were found in terms of functional outcome at 3 months, intra-hospital death, and mortality at 3 months (Supplementary Table 2).

### Plasma biomarkers

Patients with ischemic lesions at T1 showed a higher median plasma concentration of NOX 2 at T0 compared to those without ischemic lesions, although the difference had a borderline statistical significance (34.9 pg/ml vs 22.4 pg/ml, p=0.051). (Supplementary Table 3 and Supplementary Figure 1). For the remaining serum biomarkers, no statistically significant differences were found between the two groups at the three time points (Supplementary Table 3 and Supplementary Figure 1).

In the exploratory multivariate analysis, after adjustment for age, sex, neutrophil absolute number at T0 and hematoma volume at T1, high NOX-2 values at T0 were independently associated with the development of perihematomal ischemic lesions at T1, although with a trend toward the statistical significance (OR 1.122, 95% CI 0.991-1.269, p: 0.068).

Changes of biomarker levels from a timepoint to another in the two groups are summarized in Supplementary Table 4 and Supplementary Figure 2.

### Association between plasma biomarkers and radiological characteristics of hematoma and clinical outcome

Higher levels of ET-1 at T1 significantly correlated with smaller hematoma volumes at T2 (Spearman’s rho= −0.717, p=0.030) with only a trend toward the statistical significance for ET-1 levels at T0 (Spearman’s rho= −0.617, p=0.077) (Figure 3).

**Figure 3.**
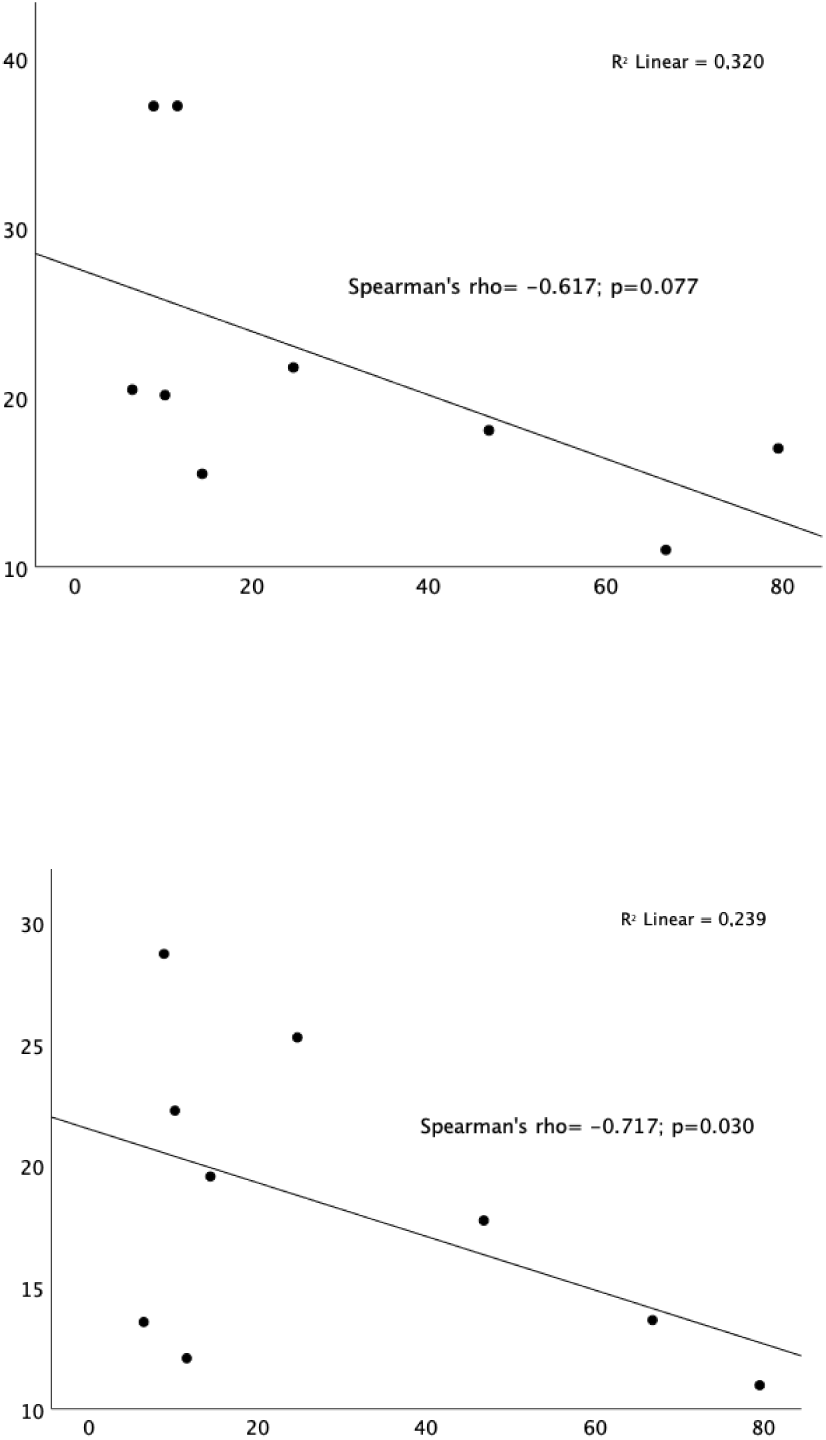
Correlations between molecular biomarkers and hematoma volume at T2.

Consistently, patients without any hematoma growth from T1 to T2 had higher levels of ET-1 at T2 compared with those with any hematoma growth (16.70 pg/ml vs. 12.83 pg/ml, p=0.020) (Figure 4).

**Figure 4.**
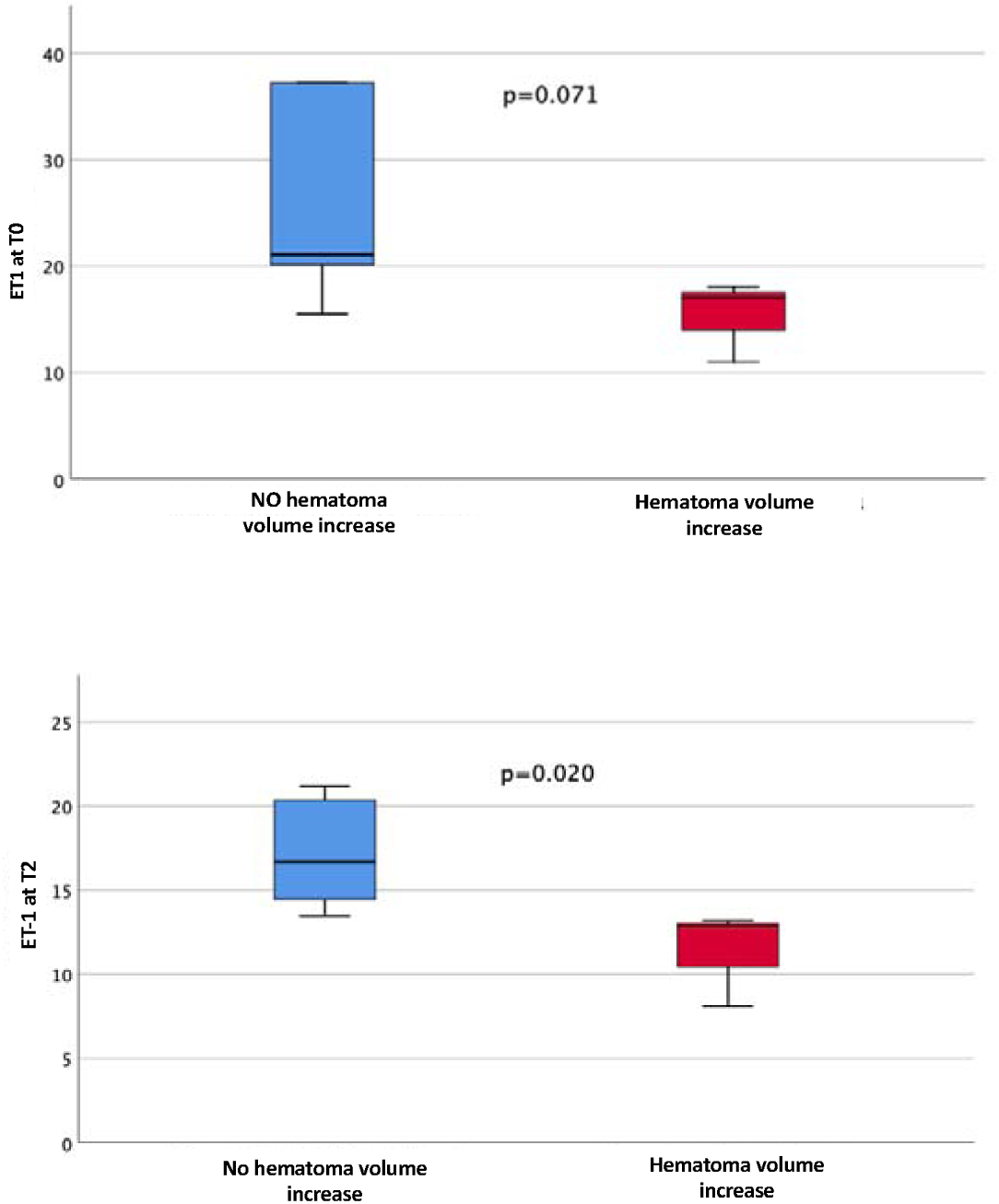
Associations between molecular biomarkers and hematoma volume increase from T1 to T2.

Higher levels of ET-1 at T1 were also associated with a borderline statistically significant absence of ipsilateral hemispheric hypoperfusion at T1 (17.76 pg/ml vs. 11.54 pg/ml, p=0.059 (Supplementary Figure 3).

Patients with a more severe perihematomal edema at T2 had higher levels of NOX2 at T0 (37.99 pg/ml vs. 19.17 pg/ml, p=0.011) and of NO at T2 (32.64 μM vs. 14.03 μM, p=0.039) (Supplementary Figure 4).

A significant linear positive correlation was found between high levels of some biomarkers and neurological severity measured by NIHSS score at different timepoints, such as NO at T1 (Spearman’s rho=0.783, p=0.007) with NIHSS at T1; MMP-12 at T1 (Spearman’s rho=0.731, p=0.040) with NIHSS at T2; while there was an inverse correlation between ADMA levels at T2 and NIHSS at T2 (Spearman’s rho= −0.743, p=0.009) (Supplementary Figure 5).

We did not find any significant association between levels of biomarkers at any timepoint and intra-hospital mortality or clinical outcome at 3 months measured by mRS score (Supplementary Figures 6 and 7). Patients with high levels of NO at T0 (28.86 vs 8.69 in the survivors, p=0.035) and lower levels of MMP-12 at T0 (875.78 vs. 1405.05, p=0.049) were more likely to die at 3 months (Supplementary Figure 8).

## Discussion

In this study, we aimed to identify biological markers potentially involved in the development of secondary brain injury following ICH, specifically focusing on perihematomal hypoperfusion, ischemic lesions, hematoma expansion, and edema development.

The pathophysiology of ICH is still not fully understood. Beyond the initial phase, where extravasated blood causes mechanical disruption of brain structures and increased intracranial pressure, a second phase follows. This phase is characterized by hematoma expansion and the development of vasogenic edema due to the rupture of the blood-brain barrier (BBB)^7^. A complex series of events occurs during this phase, involving hemoglobin, iron, hemin, and thrombin toxicity. These factors contribute to the disruption of the endothelium and tight junctions, leading to inflammatory cell invasion, increased calcium and free radical production, extensive microglia activation, and ultimately, increased neuronal death and axonal damage.^5^

Our study found that approximately 42% of ICH patients developed ischemic damage within 48-72 hours after onset, consistent with previous studies^24^. Although patients with ischemic perihematomal lesions had higher NOX-2 levels at admission compared to those without lesions, this difference did not reach statistical significance likely due to our small sample size. However, NOX-2 levels were significantly associated with the development of severe edema at 72 hours. While no significant correlation was observed between ischemic lesions and worse outcomes as measured by the mRS at three months, we did find a significant association between ischemic lesions and any increase of hematoma volume, perilesional edema, and greater midline shift. These findings suggest that oxidative stress, indicated by NOX-2 levels, plays a key role in the development of ischemic perihematomal lesions, aligning with previous research indicating that oxidative stress contributes to secondary brain injury in ICH^5–9^.

Regarding vasoactive biomarkers, our results showed an inverse correlation between ET-1 levels at 12-24 hours and hemorrhage volume at 48-72 hours. This suggests that ET-1 may exert a protective effect by inducing vasoconstriction of the ruptured vessel, limiting hematoma expansion. ET-1 is a powerful vasoconstrictor locally released by the endothelium at the site of vessel injury, initiating vasospasm as the first step in primary hemostasis, potentially leading to the cessation of bleeding^25^. In our study, patients with higher ET-1 plasma levels exhibited less hematoma growth and did not develop ischemic damage. This suggests that ET-1 might induce vasoconstriction in the damaged artery responsible for the bleeding, thereby reducing blood supply to the injured area and preventing hematoma expansion. This reduction in hematoma growth may protect the surrounding parenchyma and prevent the development of ischemic perihematomal lesions. These findings contradict those of Alioglu et al.^26^, who reported that higher ET-1 levels were associated with increased hematoma volume and worse prognosis. To the best of our knowledge, the study by Alioglu’s group is the only one in the literature that addresses the potential role of ET-1 in patients with ICH. Most research has focused on the role of ET-1 in ischemic stroke and subarachnoid hemorrhage. In ischemic stroke, an acute increase in ET-1 plasma levels has been associated with cerebral vasoconstriction, reduced regional blood flow and microcirculation, and larger infarct volumes ^27–29^. Additionally, animal studies on subarachnoid hemorrhage have demonstrated that ET-1 administration into cerebrospinal fluid can induce severe vasospasm^30–31^.

The discrepancy between the findings on ET-1 levels in our study and Alioglu et al.’s study likely results from a combination of factors, including differences in study design, timing and methodology of ET-1 measurement, patient populations, severity of ICH, clinical outcome measurement tools and follow-up times. A more detailed exploration of these variables in future studies could help to clarify the role of ET-1 in ICH and resolve these discrepancies. It is also possible that ET-1 exerts different actions depending on the timing of the bleeding onset. The vasoconstrictor effect may be beneficial at the beginning of the bleeding process, but it could become harmful in later phases when reduced blood flow might cause surrounding hypoperfusion and perihematomal damage. Moreover, the endothelin system is very complex, and its effects can vary depending on the receptors activated: stimulation of the ET_A_ receptor induces potent and prolonged vasoconstriction, inflammation, and cell proliferation, whereas stimulation of the ET_B_ receptor generally produces the opposite effects^32^.

Our analysis also identified a significant association between higher NO levels and severe cerebral edema at 48-72 hours and increased mortality at three months. It is important to note that our measurements were based on plasma nitrites/nitrates as surrogate markers for NO, which do not distinguish between NO produced by eNOS, nNOS, or iNOS. While eNOS-derived NO is generally considered neuroprotective in cerebral ischemia^33–34^ due to its vasodilatory and anti-inflammatory effects, excessive NO production from nNOS or iNOS could lead to nitrosative stress, resulting in damage to proteins, lipids, and DNA in ICH, thereby worsening outcomes. Consistent with this hypothesis, we found a significant inverse correlation between the levels of the endogenous NOS inhibitor ADMA at T2 and NIHSS scores at T2.

In the last decade, MMPs have been widely studied in both preclinical and clinical settings as potential targets for treating ischemic and hemorrhagic stroke, particularly concerning BBB disruption, inflammation, and edema development^35–36^. Numerous studies have demonstrated the deleterious roles of the gelatinases MMP-9 and MMP-2 in both animal models and humans affected by ischemic and hemorrhagic stroke^35–37^. On the contrary, no clinical studies have so far investigated the time course of the metalloelastase MMP-12 after ICH. In animal models of ICH, MMP-12 expression in the brain increases dramatically in the perihematomal zone and its suppression reduces brain damage and promotes neurological, sensorimotor and cognitive functional outcomes^38^.

We did not find any associations between higher levels of MMP-12 and increased perihematomal edema volume. Contrary to results from animal studies, MMP-12 values decrease from T0 to T2 and low levels of MMP-12 at T0 were significantly associated with a higher mortality at 3 months. These data suggest that more studies are needed to better understand the role of MMP-12 in ICH.

The primary limitation of our study is the small sample size, which may have limited our ability to detect statistically significant differences for some biomarkers. Additionally, the complexity of the study design and the use of surrogate markers for NO production could affect the interpretation of our findings.

The strengths of this study lie in its comprehensive biomarker analysis, integration of advanced imaging techniques, focus on clinically relevant outcomes, exploration of understudied biomarkers, and its prospective design.

In conclusion, our study suggests that NOX-2 is linked to the development of ischemic perihematomal lesions, while ET-1 may help to limit hematoma expansion by inducing vasoconstriction of the bleeding artery. Conversely, NO appears to be associated with worse outcomes and increased mortality, likely due to its role in free radical production. We found no significant correlation between edema volume and MMP-12 levels. Larger, prospective studies are needed to confirm these findings and further clarify the roles of these biomarkers in ICH, ultimately aiming to develop targeted therapeutic strategies to improve patient outcomes.

## Data Availability

The data that support the findings of this study are available from the corresponding author upon reasonable request.

## Acknowledgments

None

