## Supplemental Table1-4, Supplemental Figures 1-81- for "Secondary brain injury after parenchymal cerebral hemorrhage in humans: the role of oxidative stress and Endothelin-1": Supplementary material.pdf

### Supplementary Table 1. Radiological characteristics of hemorrhagic stroke in all study patients and by presence/absence of ischemic lesions at T1

| | **All patients** N=28 | **Ischemic lesions** n=12 | **No ischemic lesions** n=16 | **p value** |
|---|---|---|---|---|
| Angiography (%) | | | | 0.493 |
| - no | 14/18 (77.8) | 8/10 (80.0) | 6/8 (75.0) | |
| - yes | 3/18 (16.7) | 2/10 (20.0) | 1/8 (12.5) | |
| - yes, pathologic | 1/18 (5.6) | 0 | 1/8 (12.5) | |
| Hematoma side (%) | | | | 0.304 |
| - left | 14/28 (50.0) | 4 (33.3) | 10 (62.5) | |
| - right | 12/28 (42.9) | 7 (58.3) | 5 (31.3) | |
| - bilateral | 2/28 (7.1) | 1 (8.3) | 1 (6.3) | |
| Hematoma localization (%) | | | | 0.704 |
| - deep | 13/27 (48.1) | 5 (41.7) | 8/15 (53.3) | |
| - lobar | 14/27 (51.9) | 7 (58.3) | 7/15 (46.7) | |
| ***T1*** | | | | |
| Time of onset-CT at T1, h, median (IQR) | 2.3 (1.5-3.2) | 2.0 (1.4-5.7) | 2.32 (1.47-3.50) | 0.875 |
| Time of onset-MRI at T1, h, median (IQR) | 9.9 (3.3-21.8) | 14.8 (4.8-20.3) | 6.95 (3.23-22.70) | 0.947 |
| Hematoma volume at T1, median (IQR), cm3 | 11.9 (5.8-32.1) | 23.05 (13.40-41.03) | 6.65 (2.85-12.40) | **0.005** |
| Peri-hematoma edema at T1 (%) | | | | 0.131 |
| - absent | 2/28 (7.1) | 0 | 2 (12.5) | |
| - cortical sulci narrowing | 3/28 (10.7) | 0 | 3 (18.8) | |
| - ventricles asymmetry | 10/28 (35.7) | 4 (33.3) | 6 (37.5) | |
| - midline shift | 13/28. (46.4) | 8 (66.7) | 5 (31.3) | |
| Edema severity at T1 (%) | | | | 0.063 |
| - Absent/mild/moderate | 15/28 (53.6) | 4 (33.3) | 11 (68.8) | |
| - Severe | 13/28 (46.4) | 8 (66.7) | 5 (31.3) | |
| Peri-hematoma edema volume at T1, median (IQR), cm3 | 11.40 (4.13-19.33) | 15.25 (11.05-27.78) | 8.15 (3.85-12.20) | **0.046** |
| Midline shift at T1, mm, mean (SD) | 2.59 (3.89) | 4.45 (4.74) | 1.31 (2.62) | **0.036** |
| Intraventricular hemorrhage at T1 (%) | 14/28 (50.0) | 8 (66.7) | 6 (37.5) | 0.127 |
| Ischemic lesions at T1 (%) | 12/28 (42.9) | | - | - |
| - Not remote | 11/28 (39.3) | 11 (91.7) | | |
| - Remote | 1/28 (3.6) | 1 (8.3) | | |
| Number of ischemic lesions a T1 (%) | 12/28 (42.9) | 12 (100.0) | - | - |

| | | | | |
|---|---|---|---|---|
| - single<br>- multiple | 7/28 (25.0)<br>5/28 (17.9) | 7 (58.3)<br>5 (41.7) | | |
| Remote ischemic lesions at T1 (%)<br>- ipsilateral<br>- controlateral | 1/28 (3.6)<br>0<br>1/28 (3.6) | 1 (8.3)<br>0<br>1 (8.3) | - | - |
| Ischemic lesions morphology at T1 (%)<br>- perihematomal<br>- not perihematomal | <br>6/12 (50.0)<br>6/12 (50.0) | <br>6/12 (50.0)<br>6/12 (50.0) | - | - |
| Island sign at T1 (%) | 14/27 (51.9) | 10 (83.3) | 4/15 (26.7) | **0.003** |
| Hemispheric hypoperfusion at T1 (%) | 5/26 (19.2) | 4/10 (40.0) | 1 (6.3) | 0.055 |
| Fazekas Scale (%)<br>- 0 (absent/very mild)<br>- 1 (mild)<br>- 2 (moderate)<br>- 3 (severe) | <br>3 (11.1)<br>10 (37.0)<br>8 (29.6)<br>6 (22.2) | <br>1/11 (9.1)<br>4/11 (36.4)<br>2/11 (18.2)<br>4/11 (36.4) | <br>2 (12.5)<br>6 (37.5)<br>6 (37.5)<br>2 (12.5) | 0.464 |
| Fazekas Scale (%)<br>- 0-1 (absent-mild)<br>- 2-3 (moderate-severe) | <br>13/27 (48.1)<br>14/27 (51.9) | <br>5/11 (45.5)<br>6/11 (54.5) | <br>8 (50.0)<br>8 (50.0) | 0.816 |
| Microbleeds (%)<br>- absent<br>- present | <br>19/28 (67.9)<br>9/28 (32.1) | <br>7 (58.3)<br>5 (41.7) | <br>12 (75.0)<br>4 (25.0) | 0.432 |
| Number of microbleeds (%)<br>- ≤10<br>- >10 | <br>7/9 (77.8)<br>2/28 (22.8) | <br>4 (33.3)<br>1 (8.3) | <br>3 (18.8)<br>1 (6.3) | <br>0.387<br>0.835 |
| ***T2*** | | | | |
| Time of onset-MRI at T2, h, median (IQR) | 67.1 (53.5-110.3) | 59.2 (47.0-110.9) | 75.71 (53.60-110.29) | 0.753 |
| Hematoma volume at T2, mean (IQR), cm3 | 10.20 (6.8-36.5) | 19.55 (9.80-42.05) | 6.90 (6.20-27.70) | **0.043** |
| Hemorrhage volume variation from T1 to T2, mean (IQR), cm3 | -1.1 (-2.30, 3.25) | -0.65 (-4.45, 2.28) | -1.10 (-1.80, 3.70) | 0.531 |
| Hematoma volume increase (%) | 5/17 (29.4) | 2/8 (25.0) | 3/9 (33.3) | 1.0 |
| Perihematomal edema at T2 (%)<br>- absent<br>- cerebral sulci<br>- ventricles asymmetry<br>- midline shift | <br>2/18 (11.1)<br>1/18 (5.6)<br>6/18 (33.3)<br>9/18 (50.0) | <br>0<br>0<br>1/8 (12.5)<br>7/8 (87.5) | <br>2/10 (20.0)<br>1/10 (10.0)<br>5/10 (50.0)<br>2/10 (20.0) | **0.040**<br>-<br>-<br>0.103<br>**0.006** |
| Edema severity at T2 (%)<br>- Absent/mild/severe<br>- Severe | <br>9/18 (50.0)<br>9/18 (50.0) | <br>1/8 (12.5)<br>7/8 (87.5) | <br>8/10 (80.0)<br>2/10 (20.0) | **0.015** |
| Perihematomal edema volume at T2, mean (IQR), cm3 | 23.50 (13.7-38.5) | 38.0 (20.18-43.08) | 14.80 (9.85-23.65) | **0.012** |

| | | | | |
|---|---|---|---|---|
| Edema volume variation from T1 to T2, median (IQR), cm3 | 4.8 (1.2, 15.4) | 12.50 (-2.03, 19.0) | 4.3 (3.10, 13.20) | 0.596 |
| Edema volume increase (%) | 13/16 (81.3%) | 5/7 (71.4) | 8/9 (88.9) | 0.550 |
| Midline shift at T2, mm, mean (SD) | 2.94 (3.54) | 4.75 (3.22) | 1.3 (2.99) | **0.035** |
| Intraventricular hemorrhage at T2 (%) | 6/17 (35.3) | 3/7 (42.9) | 3/10 (30.0) | 0.644 |
| Ischemic lesions at T2 (%) | 9/15 (60.0) | 6/7 (85.7) | 3/8 (37.5) | 0.066 |
| - Not remote | 7/15 (46.7) | 4/7 (57.1) | 3/8 (37.5) | 0.462 |
| - Remote | 2/15 | 2/7 (28.6) | 0 | 0.200 |
| Number of ischemic lesions at T2 (%) | 9/15 (60.0) | 6/7 (85.6) | 3/8 (37.5) | 0.066 |
| - single | 3/15 (20.0) | 2/7 (28.6) | 1/8 (12.5) | 0.453 |
| - multiple | 6/15 (40.0) | 4/7 (57.1) | 2/8 (25.0) | 0.220 |
| Remote ischemic lesions at T2 (%) | 2/15 (13.3) | 2/7 (28.6) | 0 | 0.200 |
| - ipsilateral | 2/15 (13.3) | 2/7 (28.6) | | |
| - controlateral | 0 | 0 | | |
| New ischemic lesions at T2 vs at T1 (%) | 3/16 (18.8) | 0 | 3/8 (37.5) | 0.200 |
| Ischemic lesions at T1 not visible at T2 (%) | 2/16 (12.5) | 2/8 (25.0) | 0 | 0.467 |

IQR= interquartile range; SD= standard deviation.

**Supplementary Table 2. Clinical outcome measures in all study patients and by presence/absence of ischemic lesions at T1**

| | **All patients N=28** | **Ischemic lesions n=12** | **No ischemic lesions n=16** | **p value** |
|---|---|---|---|---|
| mRS at 3 months (%) | | | | 0.082 |
| - 0 | 2/23 (8.7) | 2/8 (25.0) | 0 | |
| - 1 | 2/23 (8.7) | 0 | 2/14 (14.3) | |
| - 2 | 4/23 (17.4) | 0 | 4/14 (28.6) | |
| - 3 | 2/23 (8.7) | 1/8 (12.5) | 1/14 (7.1) | |
| - 4 | 2/23 (8.7) | 2/8 (25.0) | 0 | |
| - 5 | 1/23 (4.3) | 0 | 1/14 (7.1) | |
| - 6 | 10/23 (43.5) | 3/8 (37.5) | 6/14 (42.9) | |
| mRS at 3 months 0-1 (%) | 4/23 (17.4) | 2/8 (25.0) | 2/14 (14.3) | 0.602 |
| mRS at 3 months 0-2 (%) | 8/23 (34.8) | 2/8 (25.0) | 6/14 (42.9) | 0.649 |
| mRS at 3 months 0-3 (%) | 10/23 (43.5) | 3/8 (37.5) | 7/14 (50.0) | 0.675 |
| mRS at 3 months 2-6 (%) | 19/23 (82.6) | 6/8 (75.0) | 12/14 (85.7) | 0.602 |
| mRS at 3 months 3-6 (%) | 15/23 (65.2) | 6/8 (75.0) | 8/14 (57.1) | 0.649 |
| mRS at 3 months 4-6 (%) | 13/23 (56.5) | 5/8 (62.5) | 7/14 (50.0) | 0.675 |
| Intrahospital death (%) | 10 (34.5) | 3 (25.0) | 6 (37.5) | 0.687 |

| | | | | |
|---|---|---|---|---|
| Death at 3 months (%) | 10/23 (43.5) | 3/8 (37.5) | 6/14 (42.9) | 1.0 |

mRS= modified Rankin Scale

**Supplementary Table 3. Plasma levels of molecular biomarkers at various timepoints (T0, T1, T2) in all study patients and by presence/absence of ischemic lesions at T1**

| | **All patients N=28** | **Ischemic lesions n=12** | **No ischemic lesions n=16** | **p value** |
|---|---|---|---|---|
| NO, µM, mean (IQR) | | | | |
| - T0 | 18.42 (8.68-28.62) | 15.02 (6.35-27.40) | 19.71 (9.0-30.58) | 0.386 |
| - T1 | 22.76 (19.04-42.66) | 20.69 (18.45-39.0) | 29.40 (17.03-44.69) | 0.749 |
| - T2 | 28.39 (15.35-48.34) | 28.52 (14.89-48.34) | 28.08 (16.44-65.53) | 1.0 |
| ET-1, pg/ml, mean (IQR) | | | | |
| - T0 | 18.74 (14.05-22.12) | 19.92 (17.19-33.43) | 16.40 (11.38-20.22) | 0.110 |
| - T1 | 15.71 (12.46-21.62) | 18.67 (11.81-26.15) | 13.66 (13.02-20.29) | 0.630 |
| - T2 | 13.96 (12.15-17.63) | 15.07 (11.91-17.63) | 13.65 (10.78-19.51) | 0.831 |
| NOX2, pg/ml, mean (IQR) | | | | |
| - T0 | 27.51 (18.91-37.30) | 34.94 (27.96-42.0) | 22.40 (18.37-27.75) | **0.051** |
| - T1 | 14.50 (9.41-19.79) | 17.50 (8.59-20.92) | 11.57 (9.52-17.87) | 0.749 |
| - T2 | 18.73 (12.07-24.20) | 18.99 (14.50-24.20) | 14.84 (4.34-20.43) | 0.522 |
| MMP-12, pg/ml, mean (IQR) | | | | |
| - T0 | 1143.76 (807.81-1656.84) | 1164.45 (949.31-1401.83) | 1300.29 (737.75-1979.66) | 0.722 |
| - T1 | 806.05 (428.32-923.29) | 880.48 (519.07-997.55) | 747.68 (246.04-928.46) | 0.337 |
| - T2 | 857.04 (389.90-992.98) | 706.17 (377.75-1024.28) | 857.04 (350.85-958.87) | 0.831 |
| ADMA, ng/ml, mean (IQR) | | | | |
| - T0 | 128.70 (111.73-169.43) | 167.94 (119.43-175.07) | 121.89 (103.67-146.92) | 0.131 |
| - T1 | 110.69 (98.78-120.33) | 99.44 (94.28-116.27) | 116.55 (106.67-146.20) | 0.078 |
| - T2 | 116.56 (92.06-126.77) | 116.56 (97.20-126.34) | 107.56 (91.97-144.70) | 0.831 |

ADMA= asymmetric dimethyl-arginine; ET-1= endothelin 1; MMP-12= metalloproteinase 12; NO= nitric oxide; NOX2= Nicotinamide adenine dinucleotide phosphate (NADPH) oxidase.

**Supplementary Table 4. Median changes in plasma levels of molecular biomarkers over time in all study patients and by presence/absence of ischemic lesions at T1**

| | **All patients N=28** | **Ischemic lesions n=12** | **No ischemic lesions n=16** | **p value** |
|---|---|---|---|---|
| NO, µM, mean (IQR) | | | | |

| | | | | |
|---|---|---|---|---|
| - ΔT1-T0<br>- ΔT2-T1<br>- ΔT2-T0 | 9.97 (-8.86, 20.79)<br>5.96 (-14.33, 24.90)<br>20.62 (-7.24, 29.88) | 9.54 (-1.88, 16.46)<br>9.23 (-11.21, 24.90)<br>21.13 (-21.30, 0.78) | 20.79 (-9.11, 29.27)<br>-3.82 (-18.78, 24.82)<br>11.81 (-3.72, 46.63) | 0.361<br>0.670<br>0.831 |
| ET-1, pg/ml, mean (IQR)<br>- ΔT1-T0<br>- ΔT2-T1<br>- ΔT2-T0 | -0.23 (-6.66, 2.53)<br>-2.08 (-8.28, 1.58)<br>-2.62 (-12.60, 0.84) | -3.16 (-12.63, 3.66)<br>-3.93 (-10.26, 1.58)<br>-6.89 (-21.30, 0.78) | -1.09 (-5.59, 2.27)<br>-1.05 (-6.18, 5.50)<br>-0.34 (-4.61, 1.56) | 0.631<br>0.394<br>0.136 |
| NOX2, pg/ml, mean (IQR)<br>- ΔT1-T0<br>- ΔT2-T1<br>- ΔT2-T0 | -13.32 (-19.28, -4.60)<br>-1.83 (-3.82, 16.47)<br>- 17.66 (-20.70, 0.22) | -18.56 (-23.78, -12.37)<br>-1.83 (-3.70, 16.47)<br>-18.77 (-23.12, -5.19) | -7.24 (-12.18, -2.31)<br>0.13 (-5.98, 19.21)<br>-10.90 (-18.59, 14.56) | **0.037**<br>0.831<br>0.286 |
| MMP-12, pg/ml, mean (IQR)<br>- ΔT1-T0<br>- ΔT2-T1<br>- ΔT2-T0 | -573.19 (-1094.54, -295.62)<br>5.61 (-279.27, 126.72)<br>-490.61 (-1164.14, -444.08) | -518,14 (-647.78, -247.46)<br>-29.03 (-367.64, 126.72)<br>-481.67 (-635.50, -444.08) | -1056.18 (-1739.40, -193.28)<br>29.14 (-288.68, 137.13)<br>-1166.93 (-1616.66, -183.27) | 0.200<br>0.522<br>0.286 |
| ADMA, ng/ml, mean (IQR)<br>- ΔT1-T0<br>- ΔT2-T1<br>- ΔT2-T0 | -30.72 (-45.20, -6.42)<br>-7.15 (-19.88, 31.84)<br>-37.02 (-54.56, 10.39) | -38.04 (-61.38, -21.45)<br>10.86 (-16.20, 31.84)<br>-49.86 (-58.58, 10.39) | -16.27 (-34.24, 45.25)<br>-18.46 (-25.52, 30.33)<br>-13.65 (-47.13, 22.44) | 0.150<br>0.394<br>0.286 |

ADMA= asymmetric dimethyl-arginine; ET-1= endothelin 1; MMP-12= metalloproteinase 12; NO= nitric oxide; NOX2= Nicotinamide adenine dinucleotide phosphate (NADPH) oxidase.

**Supplementary Figure 1. Median plasma levels of molecular biomarkers at different timepoints (T0, T1, T2) for presence/absence of ischemic injury at T1.**

A. NO; B. ET1; C. NOX2; D. MMP-12; E. ADMA

**A.**

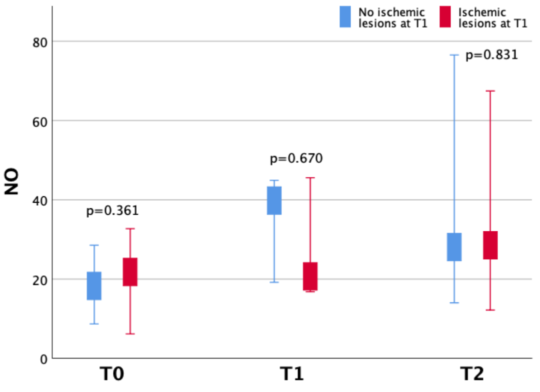


**B.**

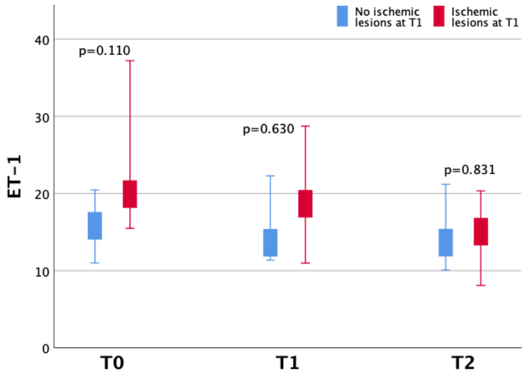


**C.**

**D.**

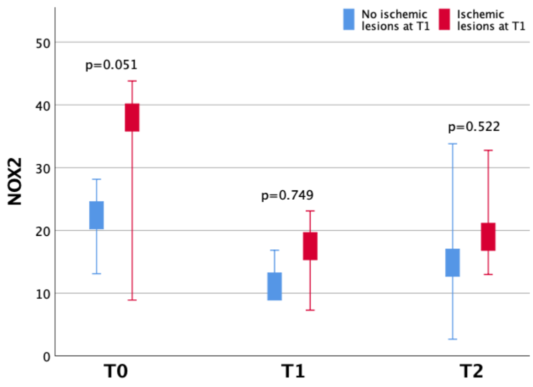


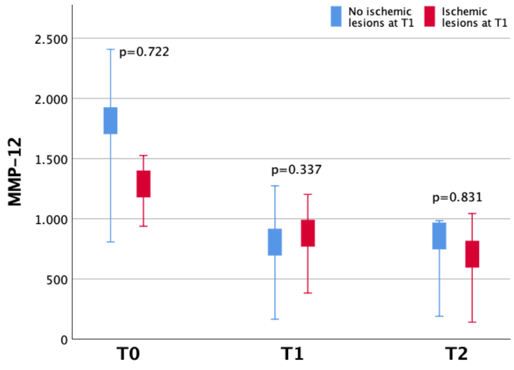


**E.**

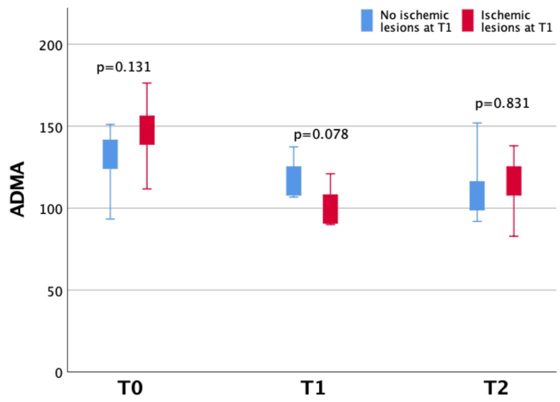


**Supplementary Figure 2. Time profile of median plasma levels of molecular biomarkers at different timepoints for presence/absence of ischemic injury at T1.**

A. NO; B. ET-1; C. NOX2; D. MMP-12; E. ADMA

**A.**

**B.**

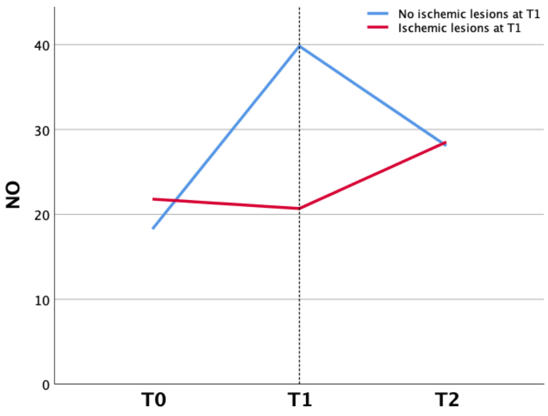


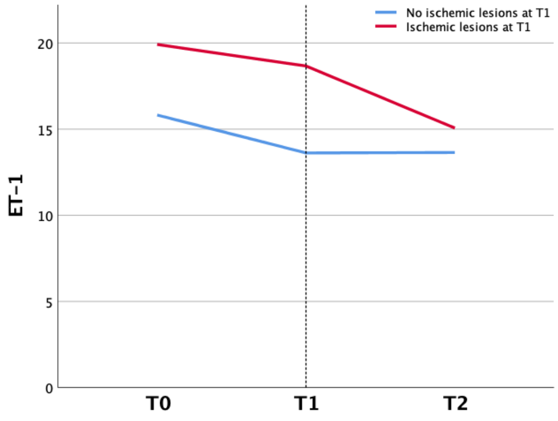


**C.**

**D.**

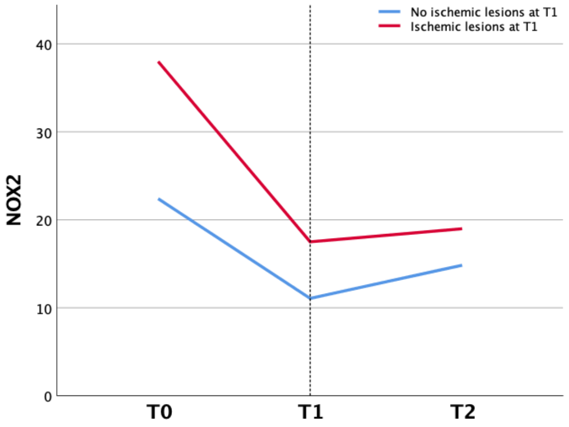


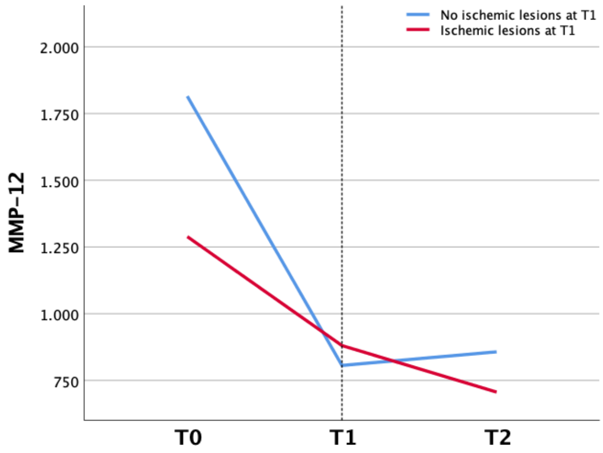


**E.**

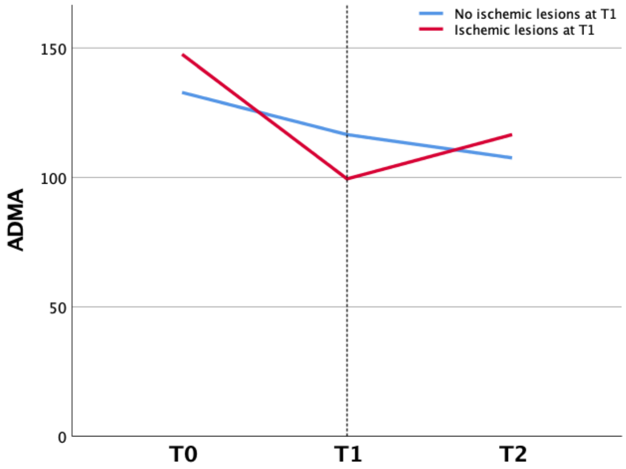


**Supplementary Figure 3. Statistically significant associations between molecular biomarkers and hemispher hypoperfusion at T1**

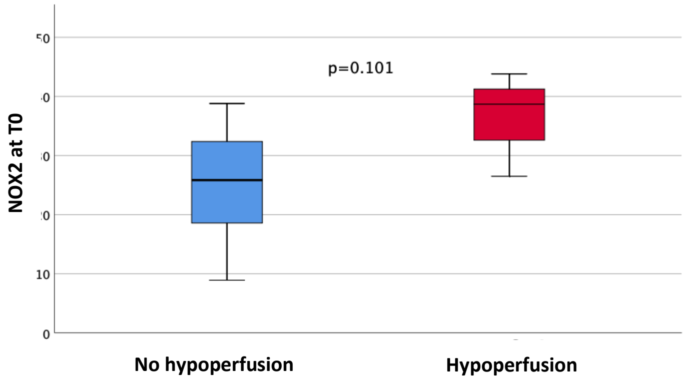


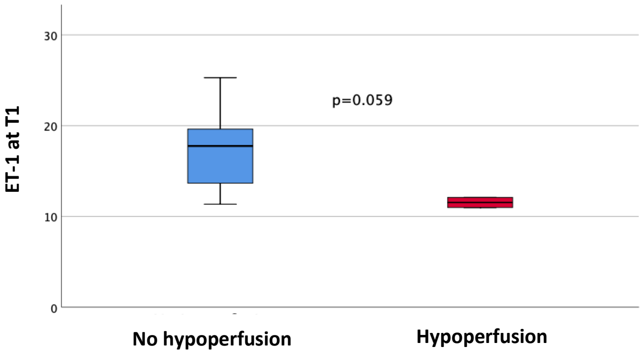

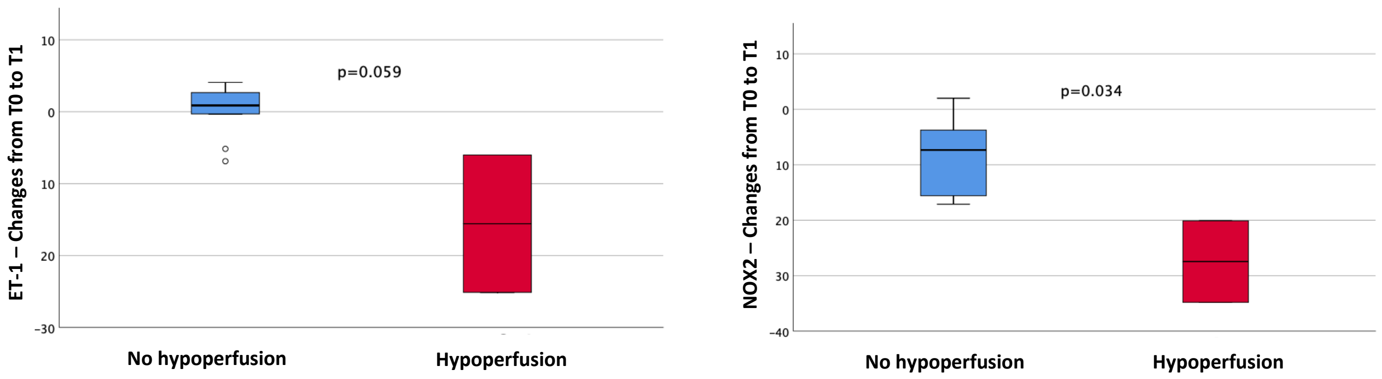


| | No hypoperfusion (n=14) | Hypoperfusion (n=3) | p value |
|---|---|---|---|
| NOX2 at T0, pg/ml, median (min, max) | 25.85 (8.91, 38.80) | 38.69 (26.49, 43.81) | 0.101 |
| ET-1 at T1, pg/ml, median (min, max) | 17.76 (11.35, 25.29) | 11.54 (10.98, 12.09) | 0.059 |
| ET-1 T0-T1, pg/ml, median (min, max) | 0.88 (-6.88, 4.08) | -15.57 (-25.12, -6.01) | 0.059 |
| NOX2 T0-T1, pg/ml, median (min, max) | -7.33 (-17.11, 2.0) | -27.45 (-34.79, -20.11) | 0.034 |

ET-1= endothelin 1; NOX2= Nicotinamide adenine dinucleotide phosphate (NADPH) oxidase.

**Supplementary Figure 4. Statistically significant associations between molecular biomarkers and severity of brain edema at T2**

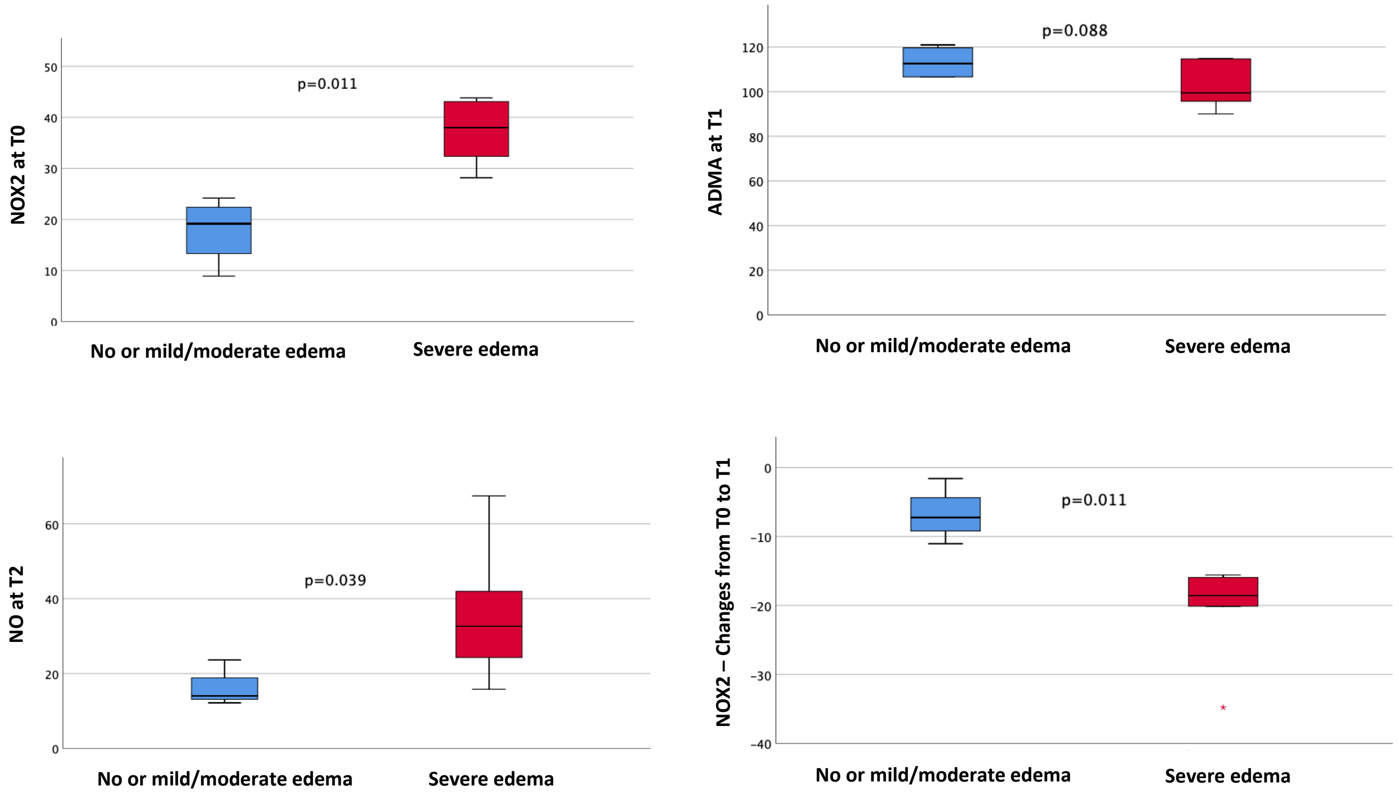

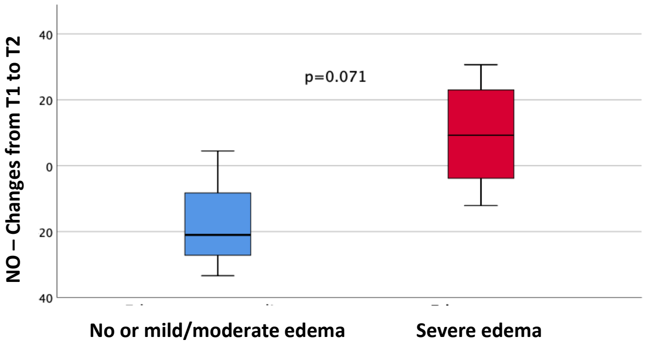


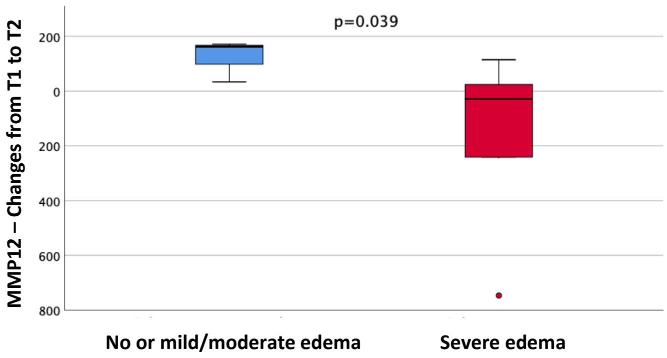


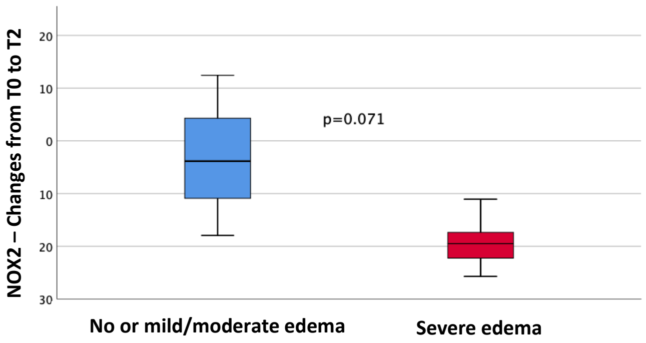


| | **No or mild-moderate edema (n=4)** | **Severe edema (n=6)** | **p value** |
|---|---|---|---|
| NOX2 at T0, pg/ml, median (min, max) | 19.17 (11.11, 23.29) | 37.99 (31.32, 43.28) | 0.011 |
| ADMA at T1, ng/ml, median (min, max) | 112.57 (106.59, 120.33) | 99.44 (94.28, 114.68) | 0.088 |
| NO at T2, μM, median (min, max) | 14.03 (12.18, …) | 32.64 (22.15, 48.34) | 0.039 |
| NOX2 T0-T1, pg/ml, median (min, max) | -7.24 (-10.12, -2.99) | -18.56 (-23.78, -15.86) | 0.011 |
| NO T1-T2, μM, median (min, max) | -21.0 (-33.38, …) | 9.23 (-5.89, 24.90) | 0.071 |
| MMP-12 T1-T2, pg/ml, median (min, max) | 162.79 (33.79, …) | -29.03 (-367.64, 47.04) | 0.039 |
| NOX2 T0-T2, pg/ml, median (min, max) | -3.12 (-17.95, …) | -19.49 (-23.12, -15.79) | 0.071 |

ADMA= asymmetric dimethyl-arginine; ET-1= endothelin 1; MMP-12= metalloproteinase 12; NO= nitric oxide; NOX2= Nicotinamide adenine dinucleotide phosphate (NADPH) oxidase.

**Supplementary Figure 5. Statistically significant correlations between molecular biomarkers and NIHSS**

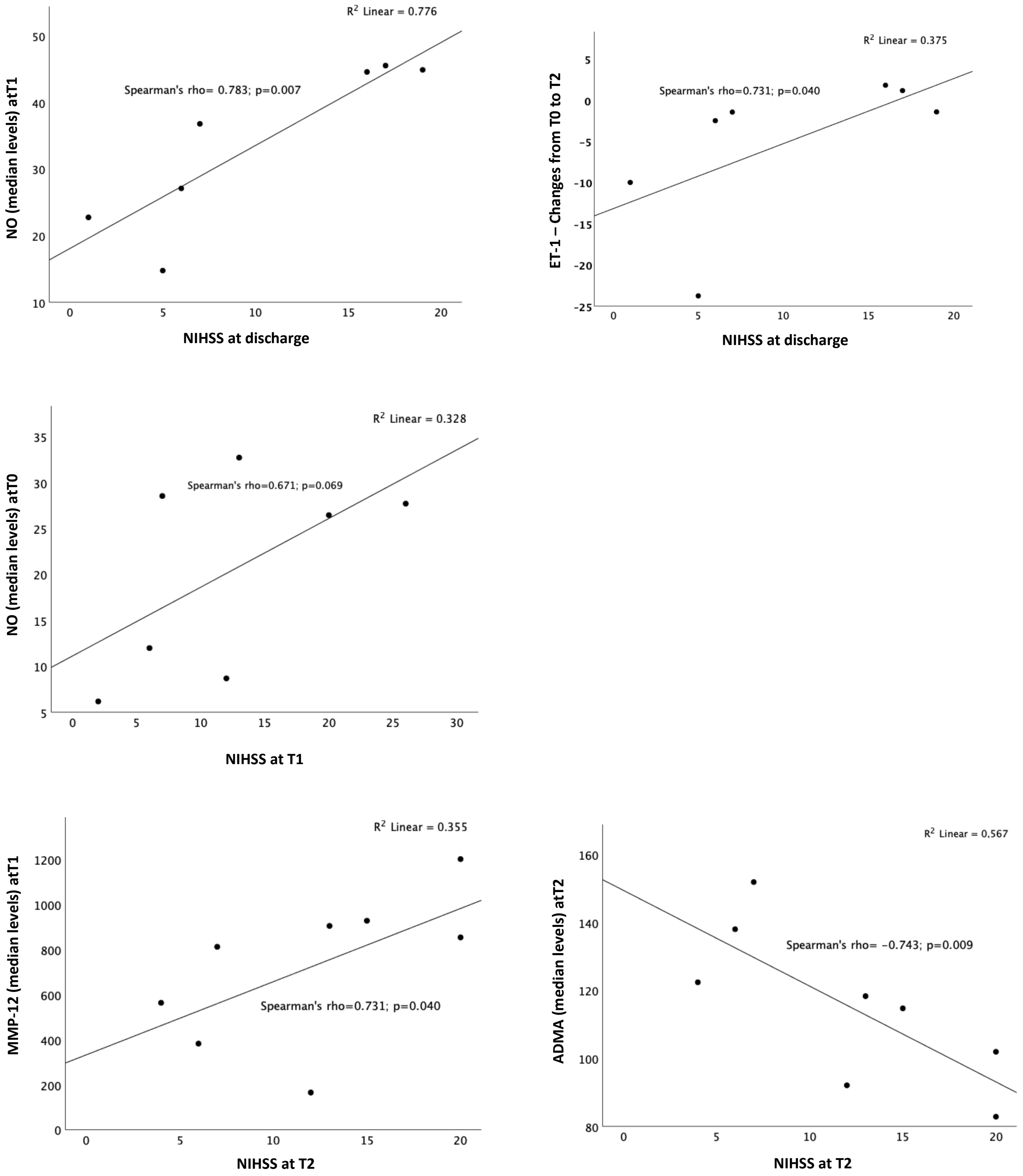

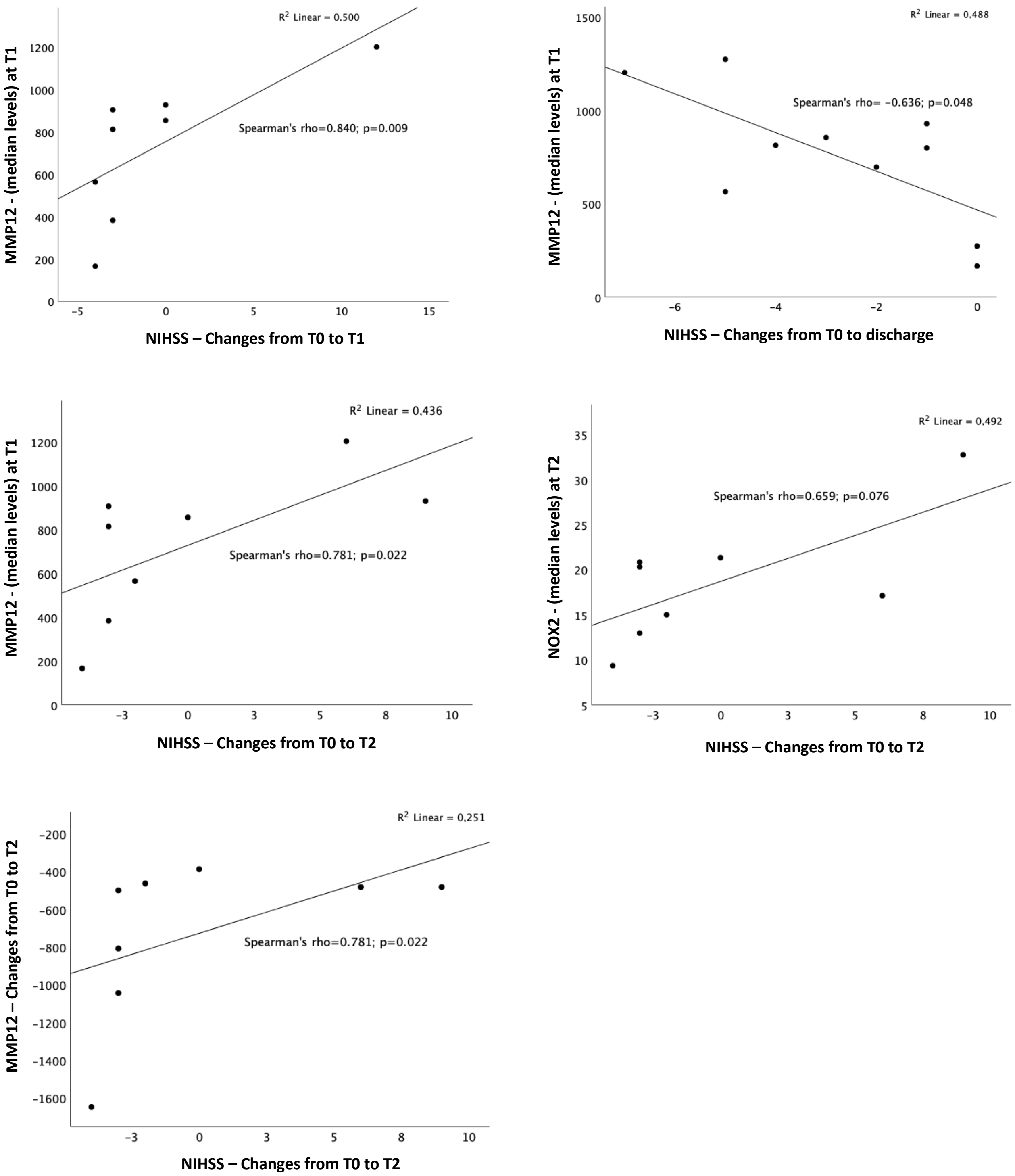
R2 Linear = 0.500
Spearman's rho=0.840; p=0.009
MMP12 - (median levels) at T1
NIHSS – Changes from T0 to T1
R2 Linear = 0.488
Spearman's rho= −0.636; p=0.048
MMP12 - (median levels) at T1
NIHSS – Changes from T0 to discharge
R2 Linear = 0,436
Spearman's rho=0.781; p=0.022
MMP12 - (median levels) at T1
NIHSS – Changes from T0 to T2
R2 Linear = 0,492
Spearman's rho=0.659; p=0.076
NOX2 - (median levels) at T2
NIHSS – Changes from T0 to T2
R2 Linear = 0,251
Spearman's rho=0.781; p=0.022
MMP12 – Changes from T0 to T2
NIHSS – Changes from T0 to T2

**Supplementary Figure 6. Statistically significant associations between molecular biomarkers and clinical outcome at 3 months**

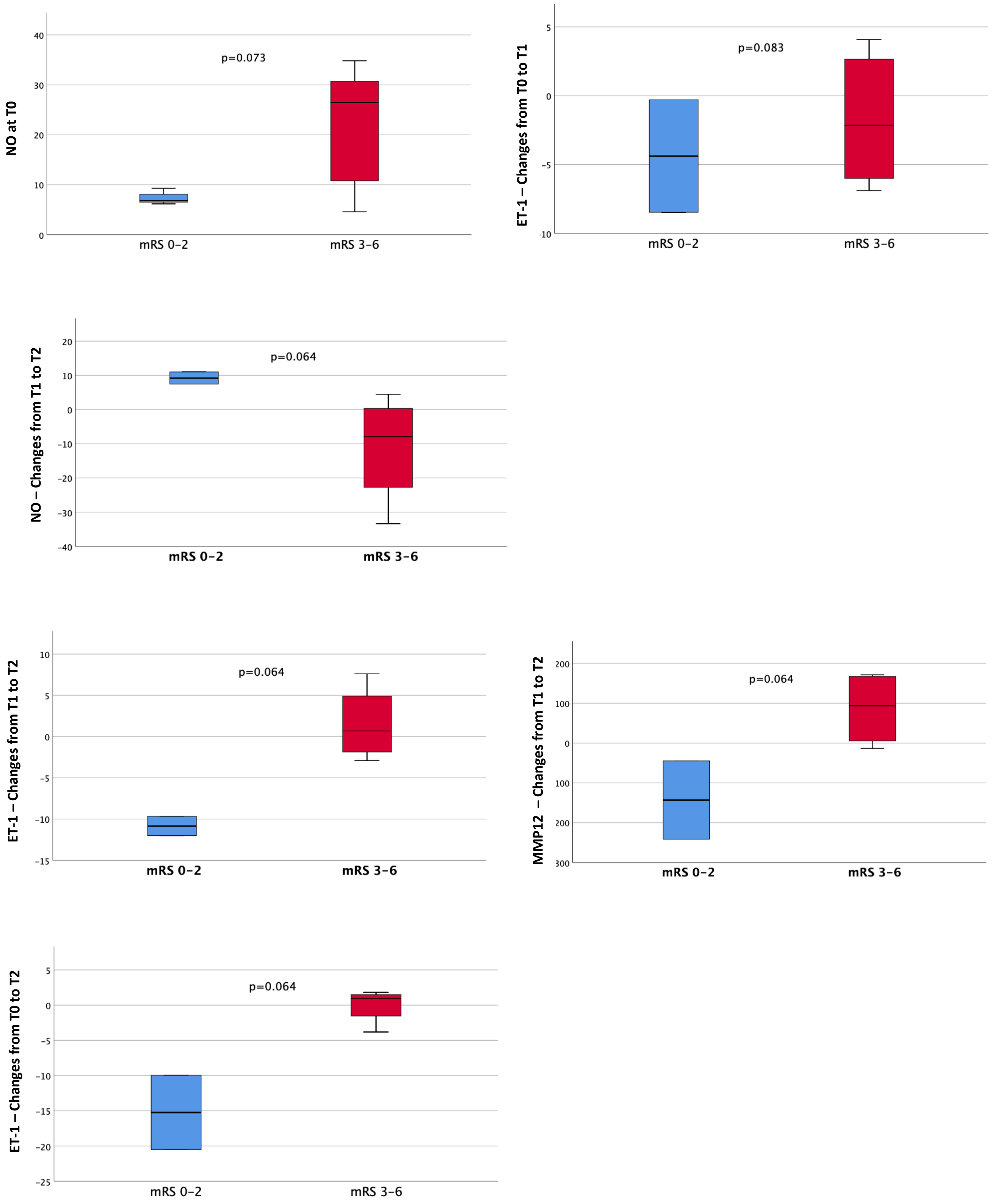

| | mRS 0-2 (n=3) | mRS 3-6 (n=3) | p value |
|---|---|---|---|
| NO at T0, median (min, max) | 6.85 (6.18, 9.31) | 26.46 (34.82, 8.69) | 0.073 |
| ET-1 at T0, pg/ml, median (min, max) | 29.17 (18.06, 37.20) | 17.38 (8.56, 34.83) | 0.083 |
| NO T1-T2, μM, median (min, max) | 9.23 (7.45, 11.01) | -7.96 (-33.38, 4.45) | 0.064 |
| ET-1 T1-T2, pg/ml, median (min, max) | -10.84 (-12.01, -9.67) | 0.69 (-2.90, 7.61) | 0.064 |
| MMP-12 T1-T2, pg/ml, median (min, max) | -143.07 (-241.34, -44.79) | 93.64 (-13.27, 171.57) | 0.064 |
| ET-1 T0-T2, pg/ml, median (min, max) | -15.23 (-20,48, -15.23) | 0.96 (-3.81, 1.83) | 0.064 |

ET-1= endothelin 1; MMP-12= metalloproteinase 12; NO= nitric oxide.

**Supplementary Figure 7. Statistically significant associations between molecular biomarkers intrahospital death**

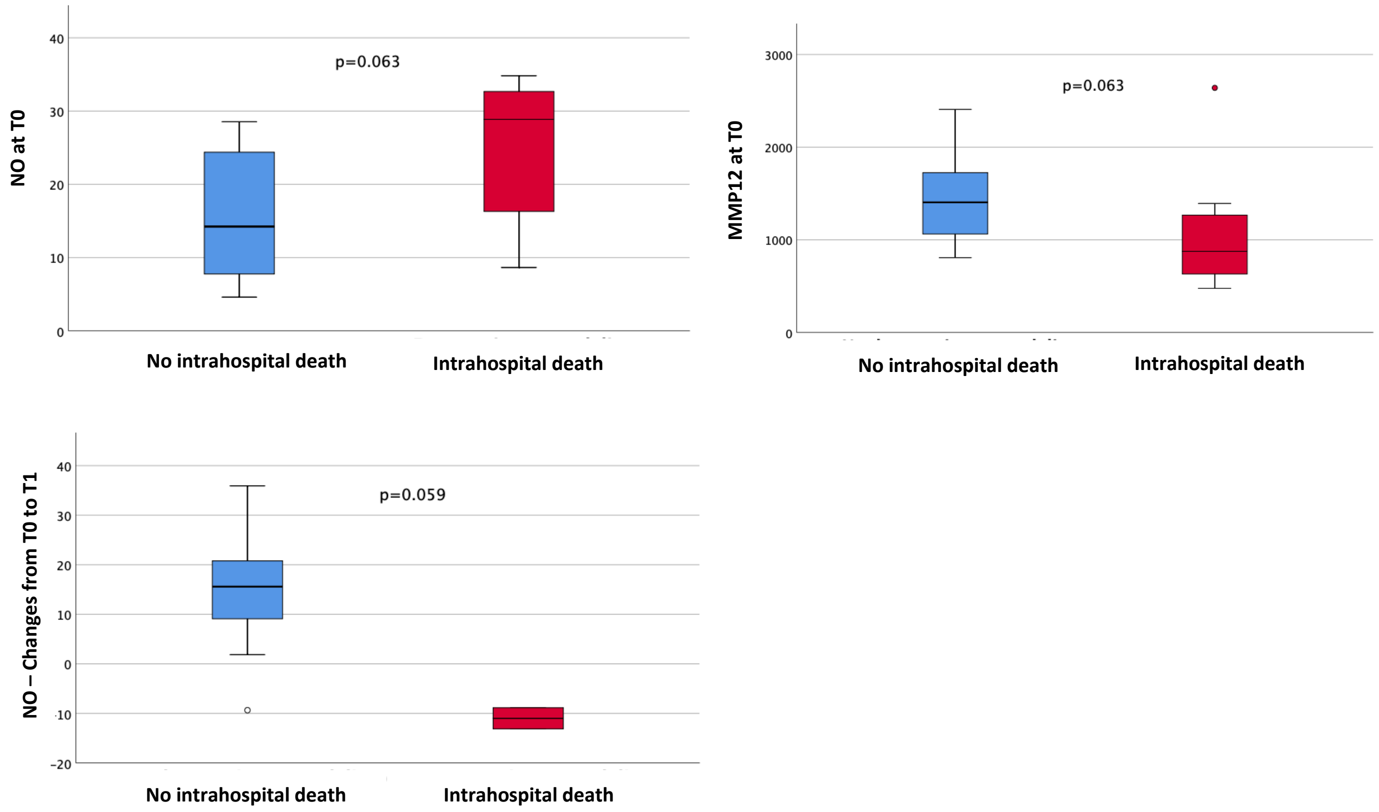


| | No intrahospital death (n=11) | Intrahospital death (n=8) | P value |
|---|---|---|---|
| NO at T0, μM, median (min, max) | 14.24 (28.54, 28.54) | 28.86 (8.65, 34.82) | 0.063 |
| MMP-12 at T0, pg/ml, median (min, max) | 1405.05 (807.81, 2407.32) | 875.78 (476.79, 2639.87) | 0.063 |
| NO T0-T1, μM, median (min, max) | 15.58 (807.81, 2407.32) | -10.99 (-13.11, -8.86) | 0.059 |

MMP-12= metalloproteinase 12; NO= nitric oxide.

**Supplementary Figure 8. Statistically significant associations between molecular biomarkers and death at 3 months**

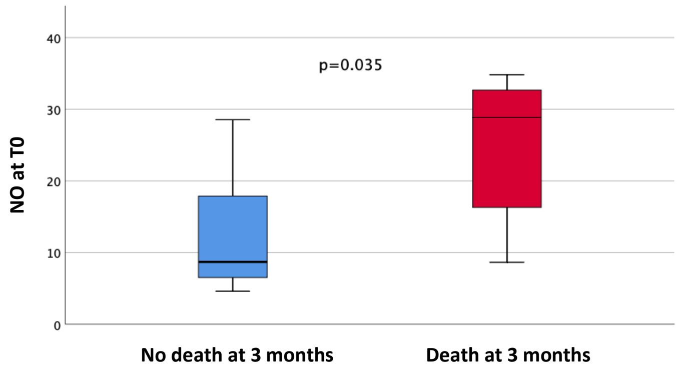


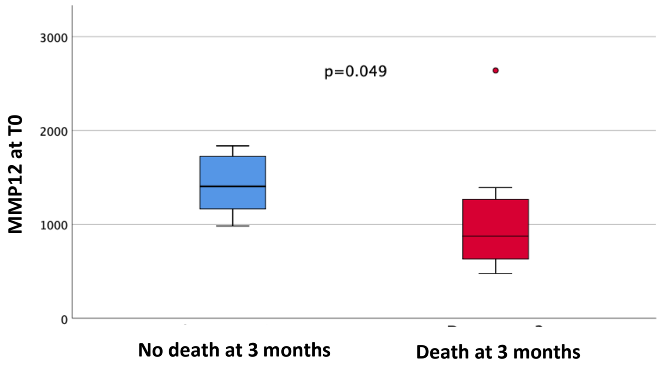


| | No death at 3 months (n=7) | Death at 3 months (n=8) | p value |
|---|---|---|---|
| NO at T0, μM, median (min, max) | 8.69 (4.61, 28.54) | 28.86 (8.65, 34.82) | 0.035 |
| MMP-12 at T0, pg/ml, median (min, max) | 1405.05 (982.66, 1837.11) | 875.78 (476.79, 2639.87) | 0.049 |

MMP-12= metalloproteinase 12; NO= nitric oxide.
